# Estimating the performance of mass testing strategies for COVID-19: a case study for Costa Rica

**DOI:** 10.1101/2022.09.05.22279618

**Authors:** Maikol Solís, Carlos Pasquier, Santiago Núñez-Corrales, Germán Madrigal-Redondo, Andrés Gatica-Arias

**Author notes:** These authors contributed equally to this work (Maikol Solís, Carlos Pasquier, Santiago Núñez-Corrales).

## Abstract

Devising effective mass testing strategies to control and suppress COVID-19 pandemic waves make up a complex sociotechnical challenge. It requires a trade-off between performing detection technologies in terms of specificity and sensitivity, and the availability and cost of individual tests per technology. Overcoming this trade-off requires first predicting the level of risk of exposure across the population available. Then selecting testing strategies that match resources to maximize positive case detection and optimize the number of tests and their total cost during sustained mass testing campaigns. In this article, we derive the behavior of four different mass testing strategies, grounded in guidelines and public health policies issued by the Costa Rican public healthcare system. We assume a (privacy-preserving) pre-classifier applied to patient data, Capable of partitioning suspected individuals into low-risk and high-risk groups. We consider the impact of three testing technologies, RT-qPCR, antigen-based testing and saliva-based testing (RT-LAMP). When available, we introduced a category of essential workers. Numerical simulation results confirm that strategies using only RT-qPCR tests cannot achieve sufficient stock capacity to provide efficient detection regardless of prevalence, sensitivity, or specificity. Strategies that harness the power of both pooling and RT-LAMP either maximize stock capacity or detection, efficiency, or both. Our work reveals that investing both in data quality and classification accuracy can improve the odds of achieving pandemic control and mitigation. Future work will concentrate, based on our findings, on constructing representative synthetic data through agent-based modeling and studying the properties of specific pre-classifiers under various scenarios.

## Introduction

The recent SARS-CoV-2 pandemic highlighted the significance of mass testing strategies as a key non-pharmacological intervention to combat and obtain accurate information about unfolding health emergencies driven by communicable diseases. Recent studies show that the effectiveness of test, trace and isolate the population depends on the ability of to estimate the current and expected pandemic impact. The task, however, needs careful evidence-based planning, avoidance of misleading metrics, and complementary information-based efforts [1]. Nations across the world must contend with a varying array of limitations in their testing and epidemiological monitoring infrastructure. Some limitations relate to systemic gaps across public health processes and procedures [2], while other manifest because of the intersection between health and economic factors leading to scarcity [3]. Sometimes testing technologies find themselves constrained under extraordinary circumstances [4].

This article departs from experiences at the intersection of limited testing capabilities with RT-qPCR, the “gold standard” [5], scarce testing alternatives, and limited information infrastructure for patient tracing. We address the question of how to maximize the effectiveness testing infrastructure with multiple test types. The study shows *in silico* the behavior of different population-level strategies under the existence of mechanisms capable of predicting individual risk of contagion. We hypothesize the population’s infection stays on *a prior* set of individual and collective factors that allow predicting the outcome. Therefore, given any available—and possibly anonymized—information, the authorities allocate testing during an emergency.

### Mass testing under pre-symptomatic and asymptomatic regimes

The recent literature has amply studies on mass testing protocols because of the challenging underlying optimization problems and the need for information for decision-making during public health emergencies. From the perspective of operations research, devising a mass testing strategy equates to solving a non-linear resource allocation problem [6]. When real-world establishes constraints, the corresponding optimization requires the use of generalized cost functions [7]. Its form, however, changes drastically between diseases with a latent or pre-symptomatic period and asymptomatic patients, and diseases without it. In particular, the testing strategy needs not keep track of prior policies adopted during the process as it has no memory. We define asymptomatic as the population infected that will never develop symptoms, while pre-symptomatic patients developing symptoms after the incubation time [8, 9].

For diseases with a latent period, we can have a closed form properties from computing optimal mass testing schedules about the ineffective process and test reliability [10]. However, the situation becomes harder for diseases with a pre-symptomatic period. Other factors alter the future outcome of the strategy: The patient’s clinical history, the disease ineffectiveness, the infection distribution, population collective behavior, geography, mobility, ongoing testing policies. Hence, the system depends on solve non-linear resource allocation problem bases on prior memory of decisions and their responses. The work by Kirkizlar et al. [11] show that, for a simple class of asymptomatic contagious diseases, determining the cost-effectiveness of dynamic intervention strategies centered on mass testing translates to a Markov Decision Process including prior data about individual test outcomes and behavioral change induced by an awareness of the disease.

Early in the COVID-19 pandemic, it became clear that its etiology included a pre-symptomatic period [8, 9, 12]. Further studies have obtained statistically reliable data about its role in viral dispersion, including the secondary attack rate of pre-symptomatic cases [13]. This finding helps partially to explain the effectiveness of pooled testing strategies [14]. Comparative analysis of the robustness of mass testing and isolation strategies indicate that pre-symptomatic and a-symptomatic sources of COVID-19 transmission are significant. Clinical care needs to proactively integrate mass testing with technologies capable of detecting the virus early to curb the disease spread [15, 16]. Simulation work on a small community with high-intensity saliva-based molecular testing [17] strongly suggests that the mechanism behind the effectiveness of mass testing against COVID-19—and more generally, any infectious disease with pre-symptomatic and asymptomatic populations—corresponds precisely to detect and isolate large share these individuals. This is the main line of defense, before the availability of vaccines to decrease the *R*_*t*_. Detailed numerical models confirm that these two populations contribute most to spontaneous transmissions, making control by non-pharmacological intervention a challenging task [18]. Mass testing at sufficient intensity increases the reliability of *R*_*t*_ for decision-making since its representativeness of the situation is inversely proportional to the reported positivity rate. It is worth remembering that the positivity rate is a function of the number of tests and the sampling strategy used, and that it does not account for unobservable variations introduced by pre- and asymptomatic cases [19].

### COVID-19 testing technologies

SARS-CoV-2 rapidly became a global threat since the first cases in China in December 2019 [20]. It prompted an arms race to achieve reliable testing at scale [21]. The world has experienced catastrophic social, economic and public health consequences. The healthcare systems overwhelmed their capacity with infected patients in their centers exceeding available capacity across multiple waves, putting a strain on the available resources and generating excess deaths from other conditions due to the saturation of patient services [22]. All nations since 2020 have implemented numerous of public health interventions, such as full lock-downs, mask wearing and social distancing, among others [23], and their effects were recently found to be synergistic [24]. Even in the context of vaccination, nonetheless, mass testing remains a powerful tool to contain, mitigate and control the spread of the pandemic at lower social, economic and individual costs compared to other interventions.

It became clear during the first stage of the pandemic that mass testing constitutes a key policy measure in the context of rapid contagion across borders [25], driven by asymptomatic and pre-symptomatic individuals. Studies revealed a conservative estimate of 30–45% asymptomatic cases, the latter figure including pre-symptomatic ones. Iceland and Indiana (randomly selected individuals), Vo’ Italy (almost all residents), the repatriate Greek persons evacuated from the UK, Spain and Turkey or Latino workers in San Francisco [26–28] exemplify some locations where RT-qPCR backed mass testings calculate the pre- and asymptomatic portion of the population. Simulation models strongly establish that these two populations drive a large share—if not the largest—of disease spread dynamics in COVID-19. Therefore, the detection of silent transmission across these two populations constitutes a substantial public health target.

COVID-19 detection through a reverse transcription–quantitative polymerase chain reaction (RT-qPCR) molecular test constitutes the accepted *“gold standard”* for clinical detection thanks to their established high sensitivity and specificity (above 95%) during *in vitro* studies [29]. Despite its accuracy, RT-qPCR requires robust laboratory facilities and trained staff, both limited in number and availability.

Reporting of results normally takes 2–5 days depending, on the healthcare system capacity to process samples and perform administrative follow-up [30]. The cost per test ranges between $50 to $100 per result [31], contingent on reagent availability in global markets due to competition. Both technical requirements and market availability become obstacles to detection across the Global South [32], thereby risking sustained contagion worldwide due to international travel patterns and prompting substantial worldwide health inequalities. During high peak waves, global scarcity of reagents forces to laboratories to use alternative options [33]. Mass testing using RT-qPCR for the entire population is infeasible for most countries. Moreover, increased testing volumes can rapidly saturate a limited number of analysis facilities, leading to increased number of contagions and further spread of the disease [34].

Antigen-based tests comprise a less expensive alternative strategy to RT-qPCR. Those could cost between $30 to $50 and can give a result within 15 minutes on average [35], and a maximum of two hours [36]. Most antigen-based testing utilize microfluidics to detect either the nucleocapsid or the spike proteins [37]. Detection becomes reliable during the first week after symptoms onset [38], and *in vitro* studies show high specificity (*>*99%) but low sensitivity (*>* 66% for nucleocapsid, *>*85% spike) [37, 39]. In practice, these figures mean that the proportion of false negatives can increase to unacceptable levels when testing occurs after the week in which first symptoms appear. To provide a baseline for antigen-based alternatives, WHO established in 2020 a minimum sensitivity of 80% and specificity of 97% compared with RT-qPCR. The Center for Disease Control and Prevention (CDC) published a set of guidelines and good practices along these lines [40], including confirmatory RT-qPCR test when antigen-based alternatives yield inconclusive results.

Reverse Transcriptase Loop Mediated Isothermal Amplification (RT-LAMP) constitutes a molecular testing technology with sensitivity and specificity comparable to that of RT-qPCR. Its detection limit for coding sequences of *ORF1* and *N* ranges between 10-25 copies per *μ*L, and result times are within those of antigen-based testing [41]. In addition, RT-LAMP testing requires lower bio-safety standards (level 1 compared to level 3 for RT-qPCR). Due to a preliminary inactivation phase, the sampler sample handling can be optimized to minimize manipulation and thereby risk to others, and their interpretation is colorimetric. In general, the protocol can be scaled up, optimized [42] and distributed due to the use of abundant standard reagents [43] and, when tests are inconclusive, these be repeated at low costs. In relation to the progression of the disease, preliminary research on the sensitivity of RT-LAMP found its value on 85.2% during the first nine days of infection and 44.4% afterwards, and an average of 60% for asymptomatic patients [44]. Detailed accounts of the technique can be found in [45–50].

### COVID-19 in Costa Rica: reactive testing with RT-qPCR

In Costa Rica, COVID-19 entered the territory via air travel during early March, with the first positive case detected on March 6, 2020. During the early phase of the pandemic (March-June), RT-qPCR remained as the primary line of detection, coupled with contact tracing. Thanks to the low number of cases with respect to installed testing capacity, simulations comparing against active cases were still representative of the epidemiological reality [51]. Since contact tracing was still feasible, 7-day average positivity remained below 10%, indicating that most test were proactively driven by contact tracing.

However, from June 9, 2020, to this date, the spread of the pandemic rapidly forced testing to become reactive, and reflect the administrative reality of symptomatic patient reception in emergency services as revealed by a drastic jump in 7-day average positivity between 17% on June 10, 2020, and up to 60% on September 16, 2020. Given that testing capacity remained constant up to December 2020 and testing was reactive, RT-qPCR is unlikely to have had played a role as a significant non-pharmacological intervention. Testing capacity did not exceed 5000 daily samples (0.1% of total inhabitants). A preliminary study in 2021 [52] using MCMC simulation and wavelet analysis found statistically significant evidence that, based on the collection of official policy interventions sanctioned by the Costa Rican government, mask wearing had the largest effect in decreasing the number of hospitalizations. We note in this case that hospitalizations constitute a more reliable observable when positivity exceeds 10% of the total number of tests.

By September 28, 2020, Costa Rica introduced regulations pertaining antigen-based testing [53], where the LS-SS-012 guidelines established their use alongside RT-qPCR confirmatory test. Despite this, however, their introduction and use started only on December 26, 2021. In the meantime, RT-qPCR testing capacity increased in the private sector, reaching up to 20,000 daily tests by January 27, 2022 (4% of total population). Even with this increased capacity, however positivity remained above 10% and remains reactive.

Private healthcare providers were first authorized to perform antigen-based tests aimed at satisfying requirements during the re-opening of air travel and certifying COVID-19 positivity for sick leaves and remote work in the private health sector. A negative antigen-based test performed by these private providers does not require an RT-qPCR confirmatory test, though. This assumes implicitly that negative patients are possibly healthy, while a negative result does not entirely discard the potential infection in the patient due to the low sensitivity’s test. Only much later their open, the authorities authorized commercial import of antigen-based tests for the public. Hence, the antigen-based testing cannot was not part of a mass testing strategy in Costa Rica.

As of today, Costa Rica lacks policies, guidelines and protocols for mass testing against COVID-19. Despite mounting international evidence demonstrating their ability to control the ongoing pandemic at a lower societal and economic cost compared to other non-pharmacological interventions such as mobility restrictions. Mass testing would therefore bring a significant improvement in the ability of the public health system to decrease the probability of service saturation during high-peak weaves and hence ensure continuity of service across all health services. To do so, however, in the context of the constraints of a small emerging economy with a high economic inequality driven by a complex wage structure (*G* = 0.52) [54] requires optimizing the delicate balance between demographic accessibility, cost-effectiveness and detection capability; doing this entails compromising guided by rational means.

The most simplistic strategy of relying solely on RT-qPCR, the dominant one to this date, remains costly and impractical. Tilting the balance towards antigen-based testing leads to false security environment. In particular, because the technology leads to an increase of false negatives. This factor combined with the arising of new aggressive variants, it will rekindle the infection process by raising the number of free agents in the population.

A more balanced—and possibly more effective—approach would aim to partition the population into groups based on their risk exposure and then apply individual strategies. If risk levels are appropriately assigned to groups by predicting them via attributes of susceptible individuals in the population and the probabilistic manifestation of risk matches the capabilities of available testing technologies, then developing a mass testing strategy becomes feasible. We foresee multiple advantages to adopting this approach.

The first benefit entails the ability to perform the simultaneously largest and most informative number of tests within the population. Pooled testing strategies share this advantage, in which the statistical structure of the measurement maximizes information gains with a limited number of individual measurements [14]. The second benefit corresponds to strategically extending the reach beyond those exposed and infected into susceptible populations, thus regaining a proactive stance with a limited number of tests and testing technologies. Doing so enables an increasingly personalized mass testing meta-strategy in which the testing technology fits the nature and features of repeated risks individuals must face [55]; this has already been performed across emergency departments [56] with RT-qPCR validation. As shown by Meystre et al., the effectiveness of such predictive analytics depends strongly on achieving sufficiently high precision and recall [57]. In situations of limited information and information quality, relaxed predictive analytics can still ensure expanding into the susceptible region as desired. Based on the latter, two broad risk categories can be drawn: a high-risk group, and a low-risk group.

The high-risk group contains all symptomatic individual, their epidemiological nexus, healthcare workers and essential workers with frequent viral exposure. It includes public transportation drivers, cleaning and maintenance workers, face-to-face customer service representatives, and educators [58]. All individuals in the high-risk group have salient features that ease their identification. Some of these include economic status, access to medical services, education, and geographic location among others. Work by Escobar et al. [59] applied a Gradient Boosting Machine (GBM) to clinical and sociodemographic factors to optimize pooled testing, measured by computing the efficiency between Dorfman’s pooling and matrix pooling strategies as well as one-stage and two-stage strategies. Reported efficiency gains were significant.

In this category, given the frequency and non-contingency of their interactions, the risk for others becomes symmetrical, and correspond to the signature of superspreaders [60]. Hence, ensuring a precise diagnosis is paramount to preventing superspreading events after successful exposure and, therefore, RT-qPCR must be used. Assuming a power-law structure in the distribution of superspreaders [61], the number of test required remains approximately constant for a fixed population size and interaction structure. Were the stratified structure to broaden at the top, the national guides of antigen-based test with confirmatory RT-qPCR become the next reasonable choice.

The low-risk group comprises individuals not yet exposed to the virus and whose features do not lead to a correspondingly high probability of contagion. It constitutes the natural target for mass testing campaigns, and the largest potential gain for proactive screening of pre- and asymptomatic populations. More specifically, this group contains primarily individuals who telecommute or regularly practice social distancing most of the time. We expect of this group low prevalence with great potential susceptible to be infected. Controlling both populations would be relieved medical centers, specially in hospital beds or ICUs.

To increase detection coverage in the low-risk group, we propose using a pooling technique. It has its origin in studies in the decade of 1940 which aimed at increasing the capacity of the detection for syphilitic men [62]. The scheme divides the total number in different pools and tests each group. The negative groups declare all the individuals as negative. With the positive ones, another round of individual testing allows to detect the infected individual(s). Doing so maximizes tests that can be carried out with a reduction in time, money and chemical reagents. Other complex pooling schemes are possible as to increase the number of positive discovering [63]. Performing multiple tests to the low-risk population constitutes an alternative to pooling. This scheme implies weekly or biweekly tests to same group of individuals. The work of [64] showed that consecutive testing reduced the number of days due to sick leaves and overall transmission. Other studies have showed that frequent testing reduces the positivity rate among workers [65–67].

This work studies the statistical and mathematical mechanisms behind massive testing strategies when using a classifier to detect subjects at risk prior to testing. We explore three different strategies. Strategy 1 follows guidelines for the high-risk group ignoring the low-risk one. For the low-risk group, Strategy 2 uses a pooling technique, while Strategy 3 uses a multiple testing scheme. We formulate a probabilistic model to quantify the costs, positive captured and number of tests per person required in each strategy. To the best of our knowledge, the Costa Rican authorities have not implemented a similar study. More generally, understanding the statistical properties of testing strategies driven by classification of susceptible individuals into risk categories has the potential of maximizing the effectiveness of existing resources under constrains of nations in the Global South.

## Materials and methods

In this study, we performed an *in silico* evaluation of the behavior of different massive testing strategies preceded by a patient classification mechanism. This section describes the contextual framework of our work.

### Sensitivity, specificity, PPV and NPV

Let *D*_*P*_ be the condition of having the disease (i.e. infected) and *D*_*N*_ the condition of being no infected. The prevalence is estimated by ℙ(*D*_*P*_) such that

ℙ (*D*_*N*_) = 1 − ℙ(*D*_*P*_). Let also *N* be the total population to undergo testing. Thus, *N* × ℙ(*D*_*P*_) are the true infected and *N* × (1 − ℙ(*D*_*P*_)) the true healthy people.

Denote as 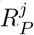 and 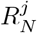 the results positive and negative of each test, respectively. In addition, let *j* = PCR, Ag or LAMP denote each available testing technology, RT-qPCR, Antigen or RT-LAMP respectively. We can thus define sensitivity the proportion of people infected who are correctly identified as positive in the test, 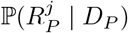. Specificity constitutes the proportion of people not infected who are correctly identified as negative in the test, or 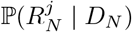.

When the prevalence is known, the relationships for testing positive or negative in a test become,

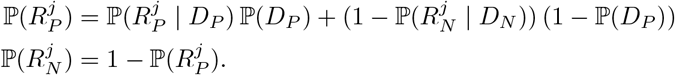

Meanwhile, the positive predicted value (PPV) is the probability of being actually positive when infected, or 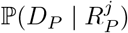. In contrast, the negative predicted value (NPV) is the probability of being negative while not having the disease, or 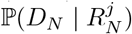. By virtue of Bayes’ theorem,

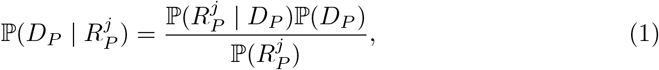

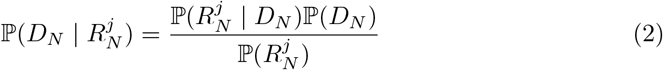

In general, sensitivity and specificity are fixed values given a testing technology. Nevertheless, NPV and PPV depend on the current prevalence. More generally, PPV increases and NPV decreases as a function of increasing prevalence. Given RT-qPCR testing has high sensitivity and specificity, PPV and NPV values are normally above 90% regardless the prevalence.

Antigen-based testing requires special attention due to its low sensitivity (80%). For a population with 25%-50% prevalence, antigen-based testing will yield a PPV of 90%-96% given that the test is used 5 days after symptom onset. Increasing prevalence, for example, above 36% yields a decrease in NPV decreases below 90%, producing many false negatives. In other words, more than 10% of tested patients were declared as negative when in reality they were infected. The procedure to follow in this case is to collect another sample across individuals whose results were negative results and perform an RT-qPCR confirmatory test.

Prevalence in the range of 1%-10% entails low impact from false negatives (NPV=98%-100%). However, an unacceptable number of false positives arises when PPV reaches values between 21%-75%. That is, large quantities of healthy people are being declared as infected when they are not, with potentially negative impacts to workforce availability. This can become particularly significant when essential workers are involved. The recommended strategy according to the [40] for this group therefore becomes to apply antigen-based testing with high sensitivity to all the population for an initial screening, and to either to perform a second round of antigen-based testing (with higher specificity) or an RT-qPCR test those whose first test was positive.

### Mass testing strategies: Pooling and multiple testing

Increasing the effectiveness of mass testing can be achieved through pooling or multiple testing. Pool testing requires three important conditions to work: (a) if all members in a group are negative then the group yields negative in the pool analysis; (b) a single positive sample within a group makes the group test result positive—further testing is necessary to identify the true positives—and (c) the fraction of expected positive cases is small. Current literature describes two large classes of pooling strategies: adaptive and non-adaptive. Adaptive ones require incremental results to further stratify testing across the population. Non-adaptive methods set the pooling scheme prior to testing, and each group is tested independent of each other. A detailed review of those techniques can be found in [63]. To simplify the estimation process, we used the most common algorithm pooling strategy, the *one-dimensional (1D) protocol*. This technique dates back to 1943 [62] when it was used to screen patients with syphilis. This scheme consists of mixing a group of samples, taken in batches. The analysis is then carried out only over these batches. If one batch is positive, then all members must be analyzed individually.

The main limitation with this technique that it becomes useful only at low prevalence levels. Consider, for example, 100 people divided into groups of 10 with only 1 positive patient (prevalence of 1%). In this situation, 9 of 10 groups will be assigned a negative result. The remainder group with one positive case should be tested entirely again. The strategy described above required 20 tests instead of 100. In contrast, if 10 people are infected and each group has one positive case in each group (prevalence of 10%), this pooling strategy results in a total of 110 tests. Other issues include loss of sensitivity due to dilution or possible artifacts introduced by the actual sample collection protocol [30, 39].

When pooling schemes are infeasible, multiple testing provides a straightforward solution. Results in [18] show that weekly testings and 2-week periods of isolation works best when transmission rates are high. If transmission rates decrease, then monthly testings and 1-week isolation periods provide the best solution to maintain the economy afloat. Re-testing strategies can reduce the sick leaves compared with no testing at all [64]. Two unpredictable factors make hard to translate results into policy. First, the asymptomatic and pre-symptomatic fractions of the population at any given time tend to be most uncertain, particularly when testing strategies are being devised; knowing how they behave explains the rate of disease spreading. Second, local transmission rates are modulated by multiple factors where little or no control is possible, including population, density, mitigation policies and local immunity [18].

The unpredictable behavior of disease transmission and the evolving response of the COVID-19 virus to survive across human populations [68] correlates well with sustained, rapid waves of transmissions. Governments continuously—yet imperfectly—have adapted to rising conditions, and then vaccines became globally available with an uneven distribution. The wide differences in testing strategies across countries, driven by limiting factors, point to the inadequacy of one-size-fits-all recommendations [69]. In this sense, remaining skeptical of the efficacy of theoretically optimal approaches appears to be essential when applied to real situational landscapes in which suboptimal strategies may be the most cost-effective ones. To ground this discussion, the following section focuses on the Costa Rican response.

### Costa Rican testing guidelines

In the Costa Rican case, the Ministry of Health in [53], defined guidelines for antigen-based testing as an alternative to RT-qPCR, depending on whether the patient is tested in public or private health services. The discriminating element is the use of RT-qPCR confirmatory test after an antigen-based test outcome is negative within the public healthcare system. The private system is excluded from required confirmatory testing. These guidelines define a *suspicious patient* (i.e., high risk) when both symptoms (e.g., high fever, cold, loss of smell sense) and a well identified epidemiological nexus (e.g., living with positive individuals, recent travel history) are present. Asymptomatic patients are deemed low risk. Therefore, the underlying principle establishes that high-risk patients must go to the public healthcare system, while the low-risk ones are directed to the private one.

Costa Rican guidelines directly follow CDC recommendations [40], which distinguish between congregate and community living setting. Congregate living entails facilities where people live in proximity to each other such as long-term care facilities, correctional and detention facilities, homeless shelters, and other group shelters. Community living is defined in contraposition to congregate living. We note that Costa Rican guidelines have failed to consider prevalence across the population as a significant factor in how they differentiate between public and private health services. The main assumption behind this, that every patient tested in the private service has a low-risk of infection and that consequently antigen-based testing is reliable, may not hold in the complex reality of disease spread of a small size, emerging economy.

### High and low risk classification

Any successful mass testing strategy should be able of to screen rapidly individuals while controlling as strictly as possible for false negatives and positives. Three elements are reported in this work to achieve this goal: cost-effectiveness, positive rate and number of tests per person. We focus on the sequence of events leading to a confirmatory test depending on whether the person is symptomatic or not and the current level of prevalence of the disease. We hence propose a set of alternative configurations informed by features of the public-private healthcare system discussed above. Our work includes a two-step strategy for massive testing: classifying the patients into high risk and low risk categories, and later applying a suitable adaptive mass testing strategy per group.

The general strategy proceeds as follows:

1. Collect or access patient data in advance corresponding to factors that determine the probability of becoming exposed to COVID-19. Due to privacy reasons or local legislation, the patient data could be confidential. In those cases, we can use aggregated statistics and estimate synthetic models to simulate a usable data table.
2. Predict patient risk categories based using data above. All symptomatic patient or those with an epidemiological nexus are automatically classified as high-risk regardless of prediction outcomes.
3. Select a strategy based on the predicted risk category:
  a. **High-risk group:** provide antigen-based testing if symptoms started 5 days or less, or provide RT-qPCR testing otherwise. All negative outcomes must be confirmed with RT-qPCR.
  b. **Low-risk group:**
    i. Use a pooling technique with a pool size of five.
    ii. Perform antigen-based testing across all groups, and perform antigen-based confirmatory testing to all members of groups with at least one positive.

The effectiveness of each strategy will depend on the prevalence, sensitivity, specificity, PPV and NPV of each test, as well as on the accuracy of the predictive model. As mentioned before, we explore only the theoretical properties of such strategies assuming an arbitrary predictive model. Models can be fitted using a wide variety of information (i.e., residence-work location, socioeconomic status, comorbidities, recent travel). Then, we will use combined characteristics of RT-qPCR and Antigen tests to create a massive strategy for all population.

For the purposes of this study, define *C*^*PCR*^ = $100 and *C*^*Ag*^ = $50 as the cost of a single RT-qPCR and antigen-based test respectively. Administrative expenses, fees and other cost were excluded. We assume a total population of *N* = 1000 individuals. For instance, using RT-qPCR test for the entire population yields *N* × *C*^*PCR*^ = $100000.

We denote a high-risk classification outcome by *M*_*H*_, and a low-risk one by *M*_*L*_. We define the classifier’s sensitivity as 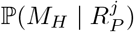, which contrasts the prediction against laboratory test results for each testing technology *j*. This value estimates the proportion of people being classified as high-risk when they have indeed a positive test result. For simplify, we assume the same sensitivity for RT-qPCR and antigen-based tests. We establish then

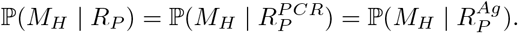

Meanwhile, the specificity 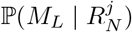 corresponds to the proportion of people classified as low-risk having a negative result. Again, we assume that both technologies have the same specificity, and we denoted just as ℙ(*M*_*L*_ | *R*_*N*_). For the purpose of our computational study, we explored classifier combinations of sensitivity and specificity at 30%, 60% and 90% for both variables.

Knowing the prevalence we can estimate

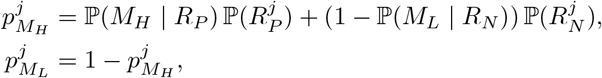

the probabilities of being classified high or low risk depending on the testing technology. To combine both probabilities, we use the logit transformation

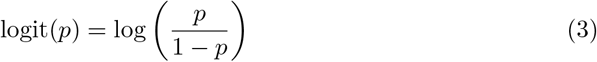

which leads to

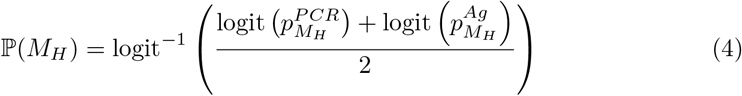

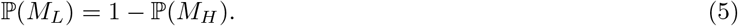

The corresponding values for PPV and NPV are 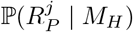 and 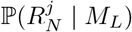.

Using Eq. 4 and 5, these become

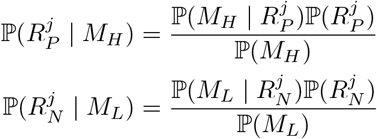

We obtain combined probabilities P(*R*_*P*_ | *M*_*H*_) and P(*R*_*N*_ | *M*_*L*_) via a similar treatment with the logit transformation.

In the context of antigen-based testing, we denote as *S*_−5_ the event of a patient has less than 5 days since the beginning of symptoms and *S*_+5_ otherwise. Since neither the high-risk condition nor the result of the test alter the distribution of patient symptoms, we assume that *S*_−5_ and *S*_+5_ are independent of 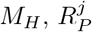 or 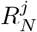. While this assumption may not hold in all the cases, it will not affect the results due to the theoretical nature of this study. To re-estimate the probability correctly when the assumption does not hold, a detailed study of the patients should make results more precise in order to confirm the hypothesis.

In our computational experiments, we set ℙ(*S*_+5_) with values of 30%, 60% and 90%. We define ℙ(*S*_−5_) = 1 − ℙ(*S*_+5_). We assume a greater proportion of RT-qPCR tests used directly on high-risk patients when ℙ(*S*_+5_) increases, and the number of antigen-based tests used at the group level increases when *P* (*S*_−5_) increases. The following sections define formulas for the overall cost, number of tests per person and number of positive reported of each strategy.

### Strategy 1: antigen-based testing

We model this scenario based on the Costa Rican public healthcare guidelines [53]. Figure 1 depicts the steps involved in this strategy, which adds a new decision layer prior to laboratory testing (blue box). The layer uses a classifier to determine high-risk (red box) or low-risk (green box) individuals. Using this label, the strategy applies different mechanism to each group. The assumptions for this scenario are:

**Fig 1.**
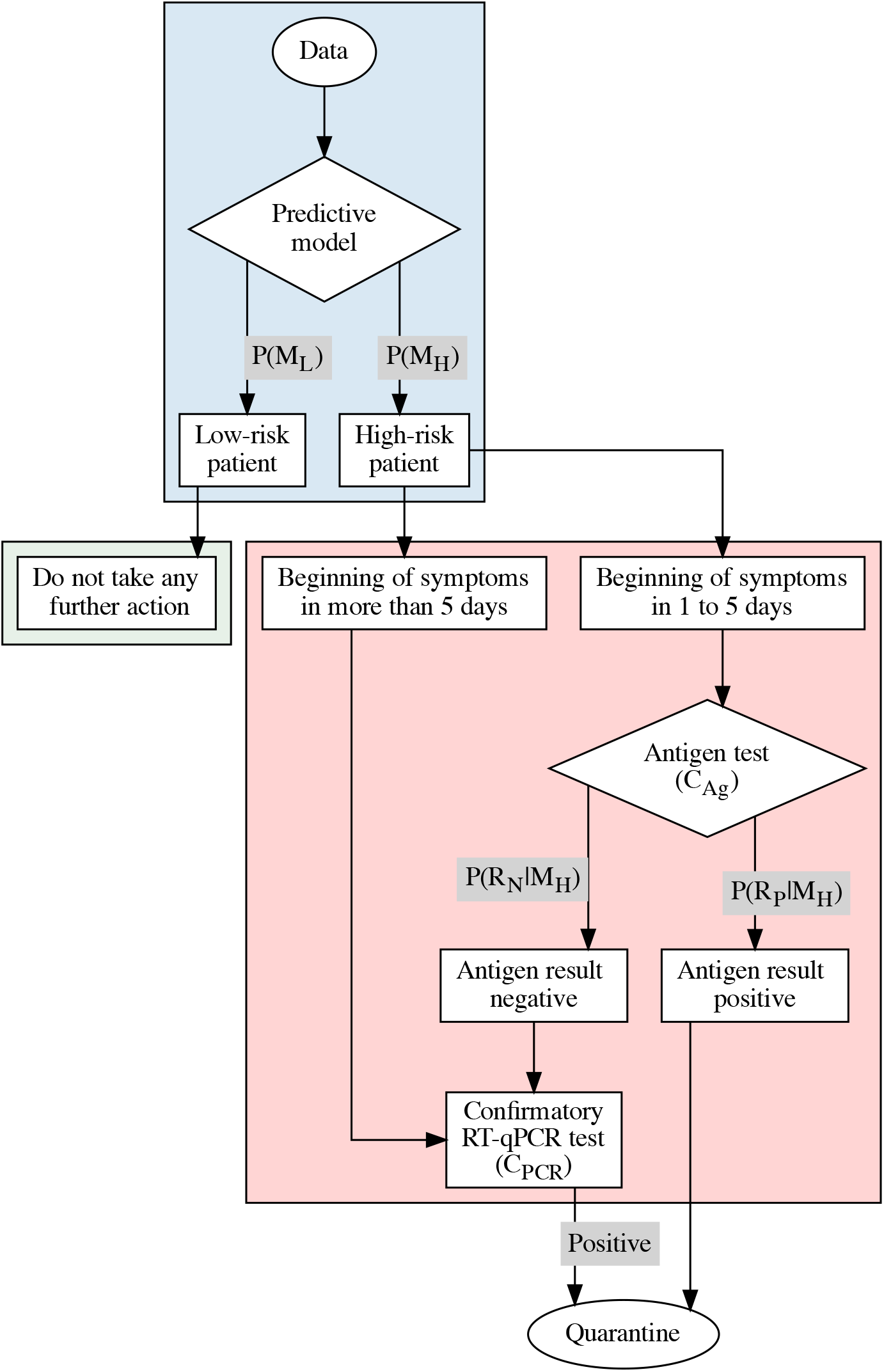
Strategy 1, mass testing with RT-qPCR and antigen-based technologies.

1. Patients in the low-risk group (*M*_*L*_) are not tested.
2. Patients in the high-risk group (*M*_*H*_) are tested according to symptom onset:
  a. Patients with less than 5 days since the beginning of symptoms (*S*_−5_) undergo antigen-based testing.
    i. If the test is positive 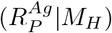, the patient is declared positive.
    ii. If the test is negative 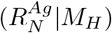, apply a confirmatory RT-qPCR (*C*^*PCR*^) test.
  b. Apply an RT-qPCR test for patients with more than 5 days after symptom onset (*S*_+5_).

To estimate overall costs, given by the number of test per person and positive captured by the strategy, we define formally each component. First, the population at risk is given by

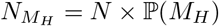

Since we only apply tests to high risk patients, we can establish the number of tests applied for each technology,

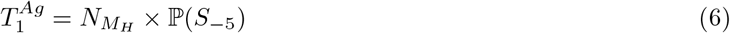

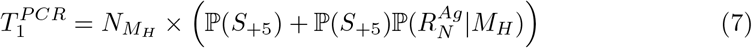

and, then, the cost for this strategy becomes

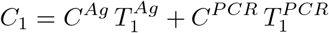

For the number of tests per person, we simply compute

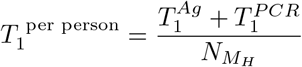

For the number of positive cases reported, we estimate the population which has undergone either antigen-based or RT-qPCR testing and multiply it by the probability of having a positive result in each case. This estimate is

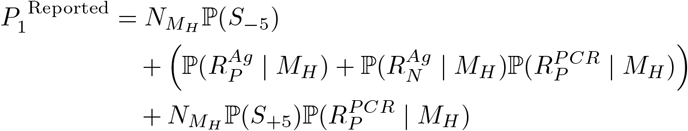

### Strategy 2: pooling

We maintain all features from Strategy 1, but include a pooling component for the low-risk group (Figure 2). The assumptions are

**Fig 2.**
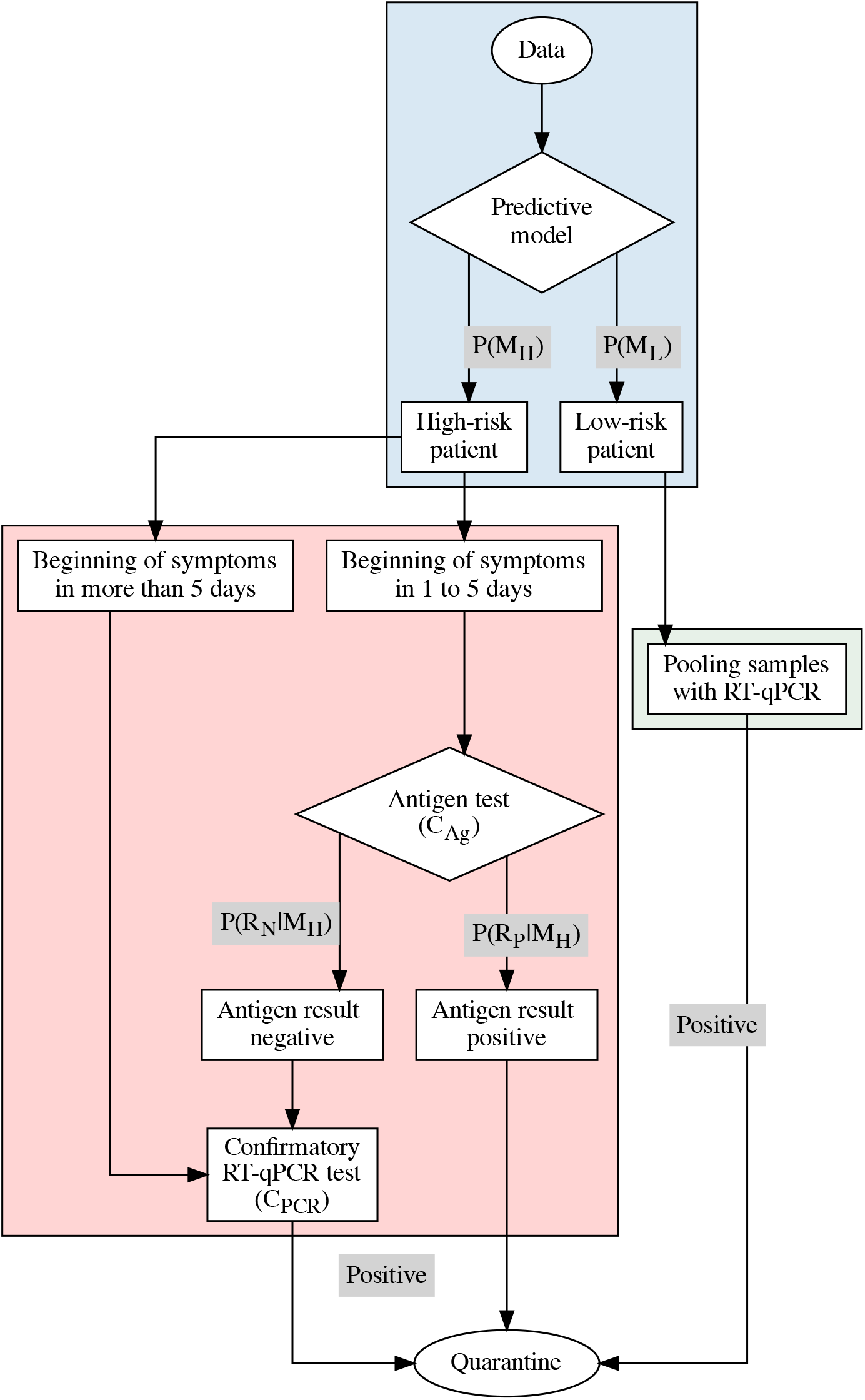
Strategy 2. Strategy 1 is supplemented with pooling for low risk patients.

1. The high-risk group follows Strategy 1
2. Pooling is applied to the low-risk group (*M*_*L*_), with pool size of 5 samples.

Similarly, as before, we define the high-risk group as *N*_*M*_ and the low-risk one as *N*_*L*_ = *N* × ℙ(*M*_*L*_). Since we did not modify the number of antigen-based tests, we use the same value 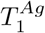 as in formula (6). The RT-qPCR tests applied in this scenario disaggregate into two components. The first one is the same in equation (7), called here 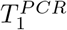. For the second one, we need to determine the number of tests used in the pooling strategy.

The first element to establish is the prevalence among the low-risk subpopulation.

Given the model, we need to estimate those individuals which are expected to be positive given the *M*_*L*_ classification. The negative predictive value of the model is given by ℙ(*R*_*N*_ | *M*_*L*_). Therefore, we define the prevalence in this subgroup as the false omission rate estimated by *p*_*L*_ = 1 − ℙ(*R*_*N*_ | *M*_*L*_).

Assuming that no loss of sensitivity occurs in the pooling technique and that the sensitivity of an RT-qPCR test is 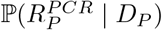, we estimate the number of positive groups

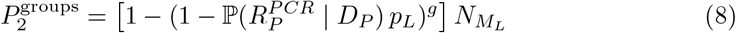

with given a total test population of size 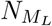 divided into groups of size *g*. The total number of test required are

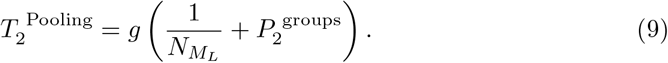

Having those elements, we define the total number of RT-qPCR tests applied as

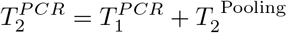

And the total cost is, therefore,

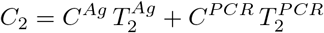

The estimate the number of tests per person required becomes

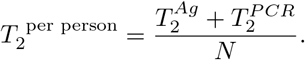

For the number of positive cases reported, we have again two components. First, we have the same number as Strategy 1 for the high-risk population. For the low-risk branch, we need to consider only those groups with positive test outcomes. We estimate the probability that their individual test in the Dorfman scheme attains a positive result. We therefore multiply the prediction outcome for pooling by the positive predictive value of an RT-qPCR test and by the group size,

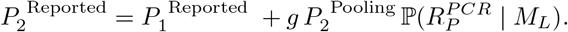

### Strategy 3: consecutive antigen-based testing

Another alternative to increase the efficiency of Strategy 1 entails applying consecutive tests to the low risk population (Figure 3). This requires applying an antigen-based test to all low-risk patients, and in case of a positive result, a second confirmatory test should be performed within the next week or two. The measure is suboptimal due to false positive rates in current antigen-based testing technologies. Confirmatory testing of positive cases prevents an increase in sick leaves or costs derived for sudden adaptation to unexpected remote work requests. However, it has been shown that even suboptimal measures were the best governments could achieve when pandemic waves emerge [18].

**Fig 3.**
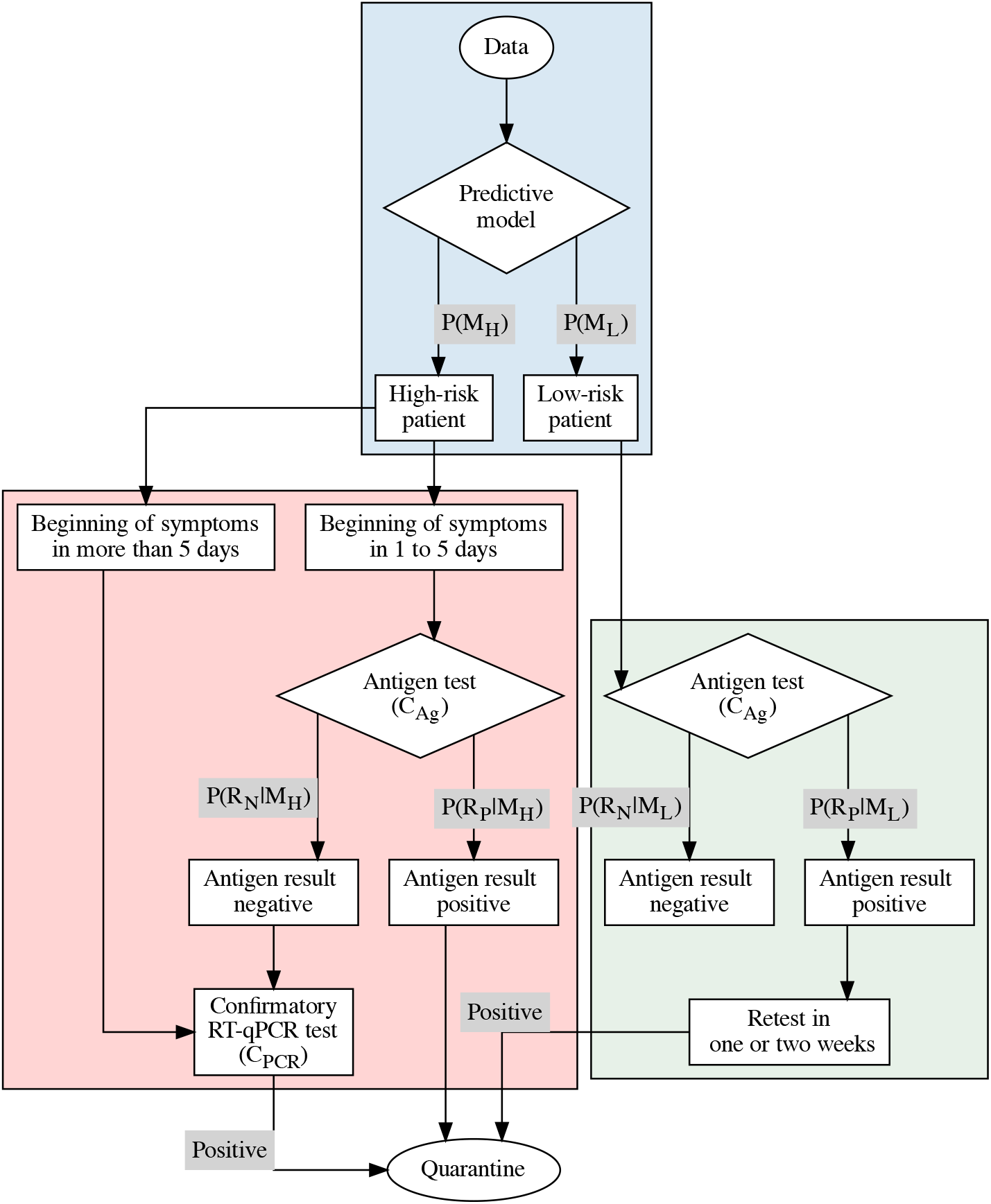
Strategy 3. Instead of pooling as in Strategy 2, all low-risk patients undergo antigen-based testing, and positive cases are required to have a similar, confirmatory test within one or two weeks.

The assumptions behind this strategy are

1. All patients in the high-risk group follows Strategy 1.
2. All patients in the low-risk group (*M*_*L*_) undergo antigen-based testing.
  a. If the result is negative, we declare the person negative.
  b. If the result is positive, we apply a confirmatory antigen-based test within one or two weeks.

The number of antigen-based test has two components due to re-testing. For the high-risk population we use the same value as Strategy 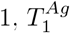. For the low-risk population, all patients undergo a first round of testing, and positive patients undergo a second one. At the end, the total number of antigen-based tests required during re-testing is

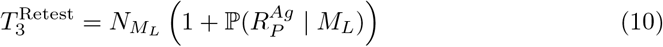

and the total number of antigen-based tests becomes

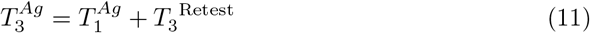

RT-qPCR tests applied are exactly the same as the Scenario 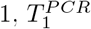. The total cost due to testing for Strategy 3 is

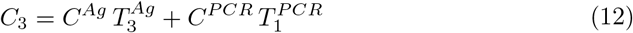

For the number of tests per person, we simply estimate

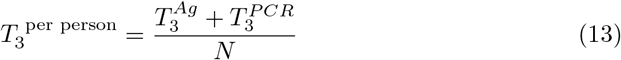

Finally, the number of positive cases reported divides into two components. First, we have the same number of positive cases as Strategy 1 for the high-risk population. For the low-risk branch, we need to consider only the test that were positives in the first or second round. This estimate is defined as

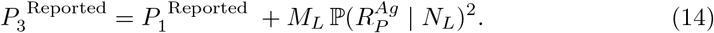

### Strategy 4: the role of saliva-based testing

Prior strategies model the current state of healthcare guidelines Costa Rica, anchored in RT-qPCR tests as the main line of defense which does not scale for mass testing purposes. Antigen-based testing has lower costs, but its low sensitivity makes confirmatory tests of negative results still necessary. An alternative solution is to include saliva-based RT-LAMP testing into the mix as suggested by a prior study [50]. RT-LAMP and other saliva-based testing technologies reach values above 90% for sensitivity and above 95% for specificity, and can be adapted quickly to new variants. In addition, the sampling process is inexpensive, requires lower biosafety standards and trained personnel than nasopharyngeal swabs. For a detailed account of a successful application of saliva-based technologies to COVID-19 prevention and mitigation, see [46, 70].

The fourth strategy proposed here seeks to overcome the flaws of other technologies by targeting them to appropriate groups based on a data-driven assessment of individual patient risk. We first separate high-risk patients further into essential workers and other high-risk. For essential workers, an RT-qPCR test is mandatory to ensure continuity of services without risking high numbers of false positives or negatives. Other high-risk patients undergo saliva-based RT-LAMP testing, well suited to in particular for high peak waves and massive screening. To capture all positive cases, a confirmatory RT-LAMP should be performed over negative cases. Finally, the low-risk group is subjected to antigen-based testing at home or in point-of-care (POC) centers. As with Strategy 3, all positive cases must confirm their result with a second test within one or two weeks.

The main assumptions behind this strategy are:

1. Essential workers are tested with RT-qPCR.
2. Patients in the high-risk group are tested with RT-LAMP.
  a. If the result is positive, we declare the person as positive.
  b. If the result is negative, we perform a confirmatory test by RT-LAMP.
3. Patients in the low-risk group (*M*_*L*_) are tested with antigen-based tests.
  a. If the result is negative, we declare the person as negative.
  b. If the result is positive, we apply a confirmatory antigen-based test in one or two weeks.

We define the new quantity

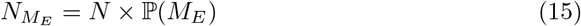

where *M*_*E*_ represents the class of essential workers on this Strategy. For simulation purposes, we set the proportion of essential workers at a fixed value of 1%. The value is a conservative estimate based on the 1.25% of total healthcare workers in Costa Rica: 2470 in the Ministry of Health, 62814 in the public social security from a total population of 5213374 inhabitants [71–73]. Therefore, we estimate the high- and low-risk groups with the remainder of the population,

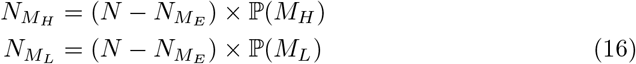

The number of tests applied in each case will depend on the technology. For RT-qPCR, we have

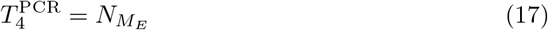

RT-LAMP tests only apply to the high-risk group, with a confirmatory test in case of negative result,

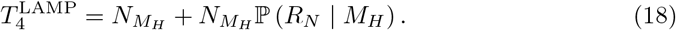

For antigen-based tests, the number is equal to that in Strategy 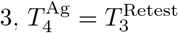 The strategy total costs become

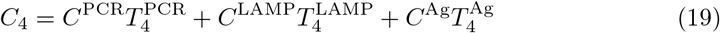

And for the number of tests per person, we estimate

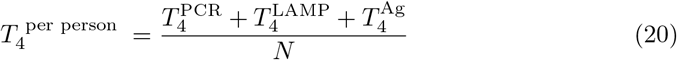

Finally, the number of positive cases can be decomposed into

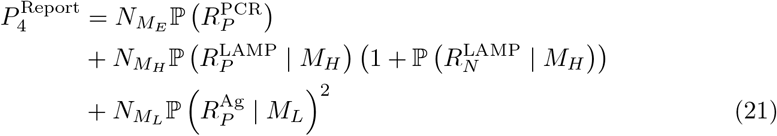

## Results

In this section, we compare the strategies above according to their costs (*C*_*i*_), number of tests per person 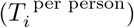 and number of positive cases reported 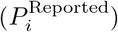 for *i* = 1, 2, 3. The total population used is *N* = 1000 and prevalence ranges from 0% to 30%. The cost of an RT-qPCR test is set to $100 and an antigen-based test to $50.

Across all figures, the red dashed line is the cost of applying an RT-qPCR test to each true infected. Formally, it is equal to 1000 × $100 × ℙ(*D*_*P*_). Those reported as positive correspond to the number of true infected individuals 100 × P(*D*_*P*_). For the number of tests per person, we set to the constant 1 indicating a baseline. Blue lines represent the percent of antigen-based tests used in each strategy according to the proportion of people showing symptoms for less than 5 days. From dark to light blue, we assume proportions of 25%, 50% and 75%. The primary *x* axis represents percent prevalence and the *y* axis varies per target: cost in dollar, number of people or tests per person. Secondary axes show the model specificity and sensitivity used in each case. Our code is available in a GitHub repository for reproducibility purposes^1^.

### Costs

Computational experiments show that using the pre-classifier reduces the total cost by correctly identifying the high-risk individuals in Strategy 1 (Figure 5). As the pre-classifier increases its predictive accuracy, cost decreases to only for those truly infected. Notice that specificity has a greater effect in reducing cost relative to sensitivity. Since this strategy excluded low-risk individuals, false negatives do not contribute to the overall cost. Conversely, false positive cases appear (i.e., false high-risk individuals), the strategy applies an antigen-based test with a confirmatory RT-qPCR in case of negative outcome.

**Fig 4.**
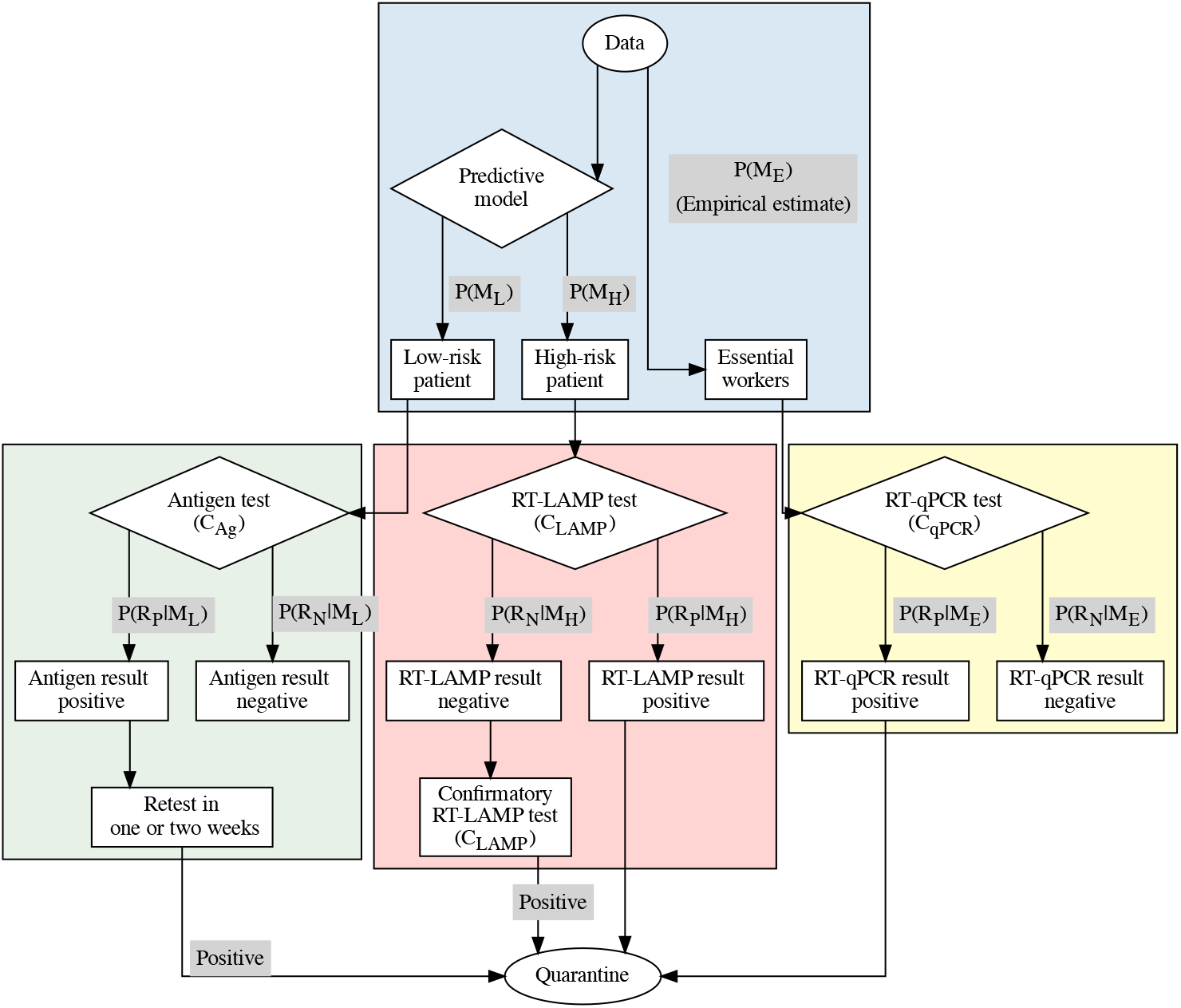
Strategy 4. The high-risk group in Strategy 3 splits into essential workers (RT-qPCR) and other high-risk patients to be tested using RT-LAMP. Low-risk patients proceed as with Strategy 3.

**Fig 5.**
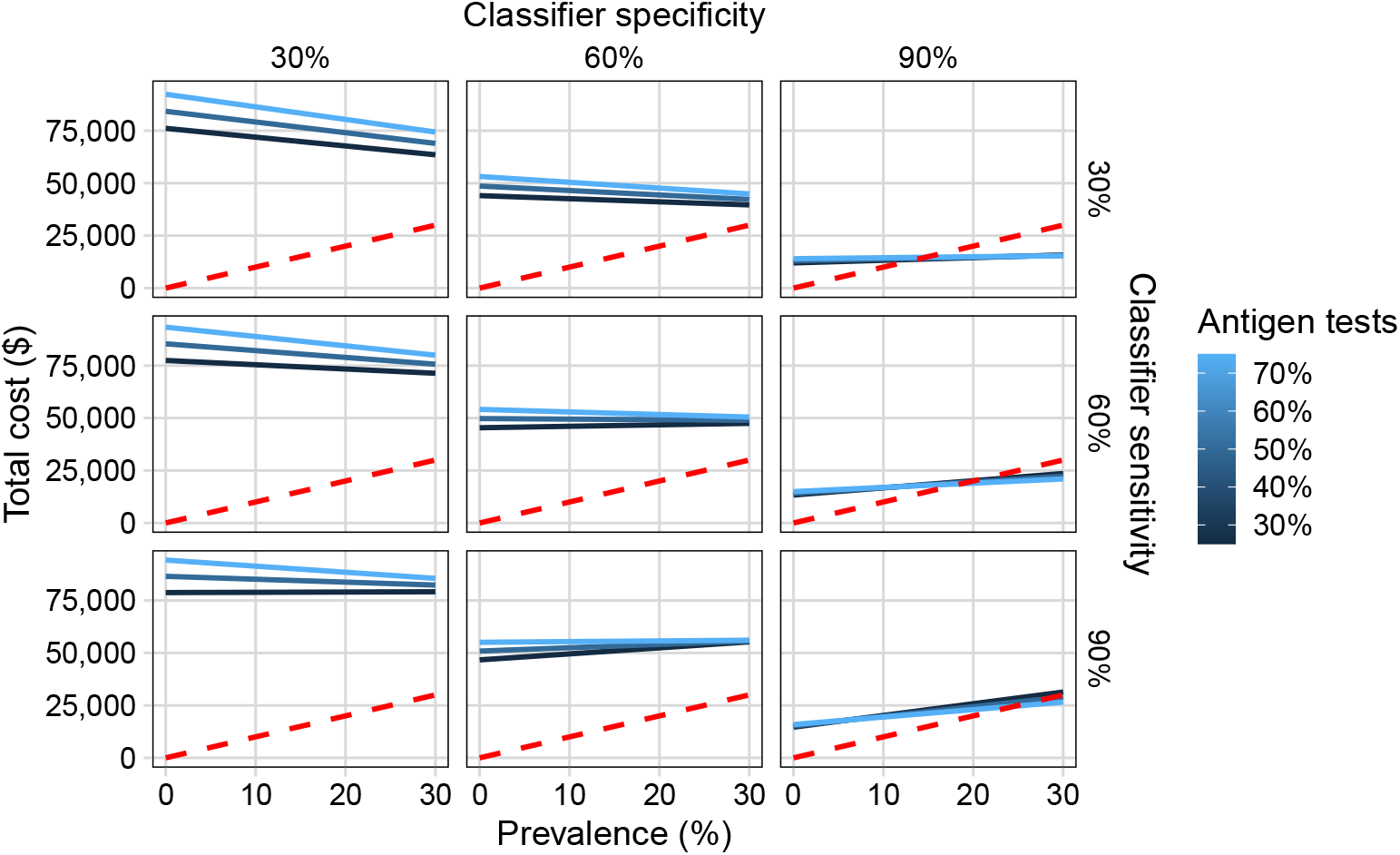
Total cost structure for Strategy 1. Using RT-qPCR tests only for the high-risk group predicted by the pre-classifier on symptomatic patients matches minimizes cost while maximizing discovery of true positives at high specificity and sensitivity values, but only for a limited portion of the population.

Sensitivity and specificity modulate the effectiveness of the classifier to rebalance the overall cost structure depending on prevalence. Specificity determines the sign of the slope of the resulting curves, while sensitivity determines the percentage of antigen-based tests applied to the population. Proportionally applying more antigen-based tests becomes more effective at prevalence values higher than 10% with tests having high specificity (90%) and medium to high sensitivity (60%, 90%).

For Strategy 2, false positive cases represent the largest cost factor (Figure 6), similar to Strategy 1. However, individuals misclassified as low-risk individuals do not increase dramatically overall costs, since it becomes a natural an overhead already accounted for in the method. Misclassifying high-risk individuals leads to incorrectly applying Strategy 1 to a healthy individual, or to applying a pooling technique to a group with at least one infected individual.

**Fig 6.**
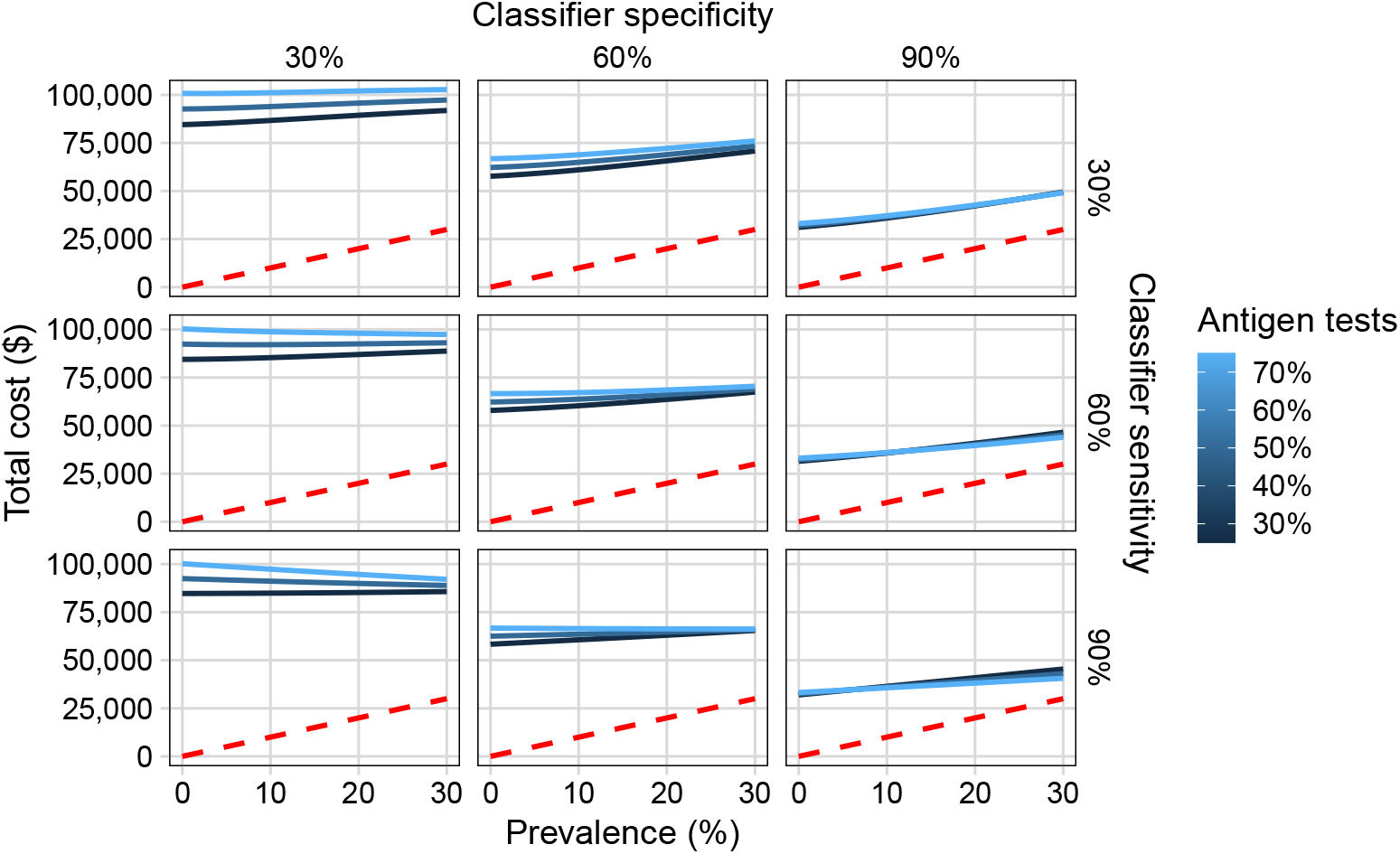
Total cost structure for Strategy 2. Pool size equals 5 samples. Cost outcomes are similar to Strategy 1 at very low prevalence, while a larger population receives testing thanks to the application of a pooling technique. Cost structure increases with prevalence, while specificity makes the effect of different antigen-based testing proportions less noticeable.

We observe how the pre-classifier helps to reduce the total cost identifying correctly the high-risk individuals. When the pre-classifier has high levels of sensitivity and specificity, we achieve outcomes similar to the Strategy 1 with a small overhead due to the cost introduced by pooling. Again, as the model becomes more accurate, this overhead decreases. Sensitivity and specificity play the same role as in Strategy 1.

In Strategy 3 (Figure 7), total costs are higher than the Strategy 1 or Strategy 2 due to massive testing with antigen-based technologies for the low-risk group. Even if it is possible to classify correctly most of the population according to their risk, the minimum will be of at least $60,000 for each 1000 individuals.

**Fig 7.**
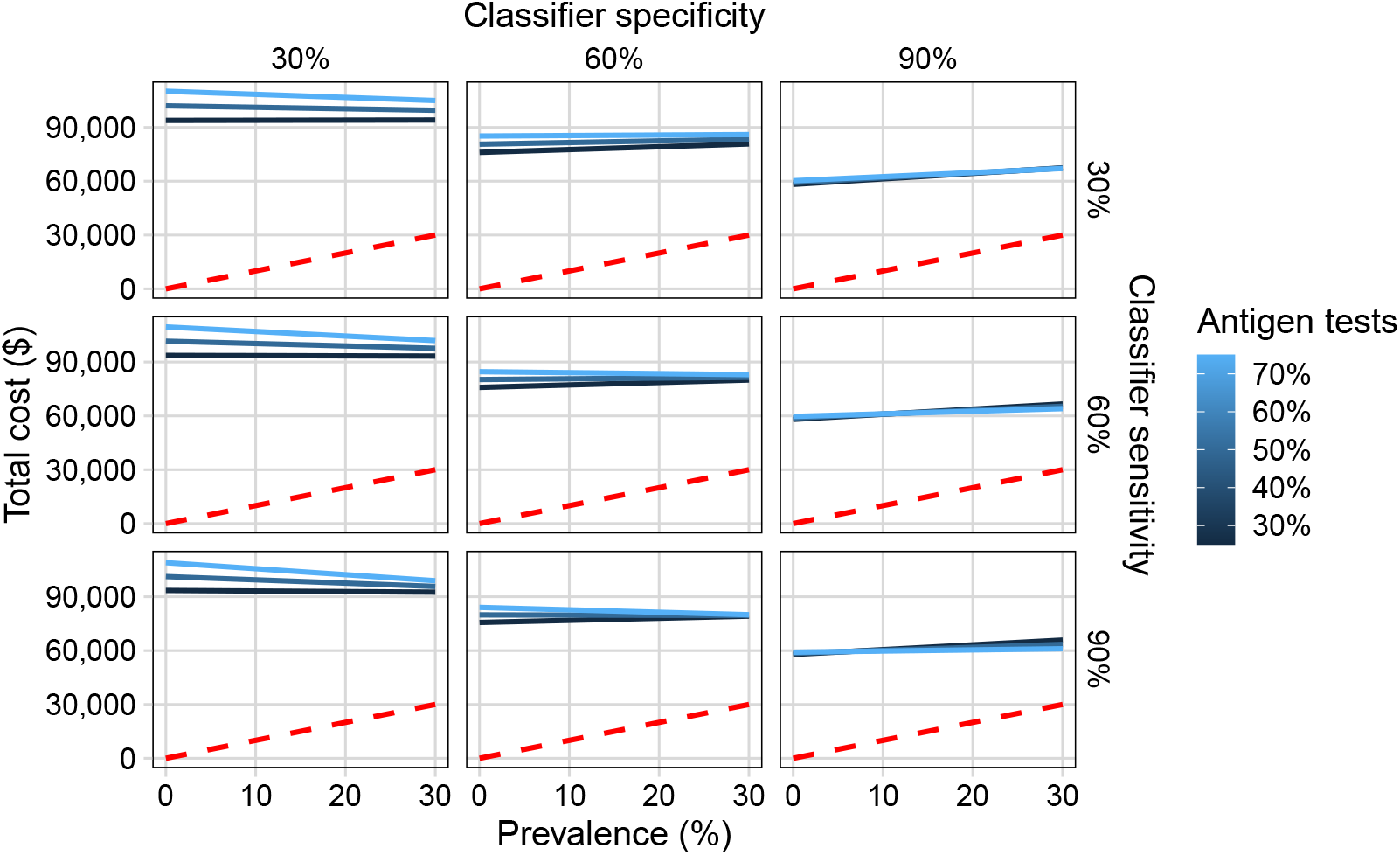
Total cost according to Strategy 3. Costs are larger than for Strategies 1 and 2 due to an increased number of antigen-based tests applied to the low-risk group. Cost structure becomes less markedly modulated by prevalence.

Finally, Strategy 4 has a similar cost structure compared with the pooling scheme in Strategy 2 (Figure 8).Using maximally targeted technologies to each type of patient is similar to applying complex (and difficult) techniques like pooling. Given that we use antigen-based testing without the restriction of incubation periods, sensitivity is the only factor affecting the sign of the slope.

**Fig 8.**
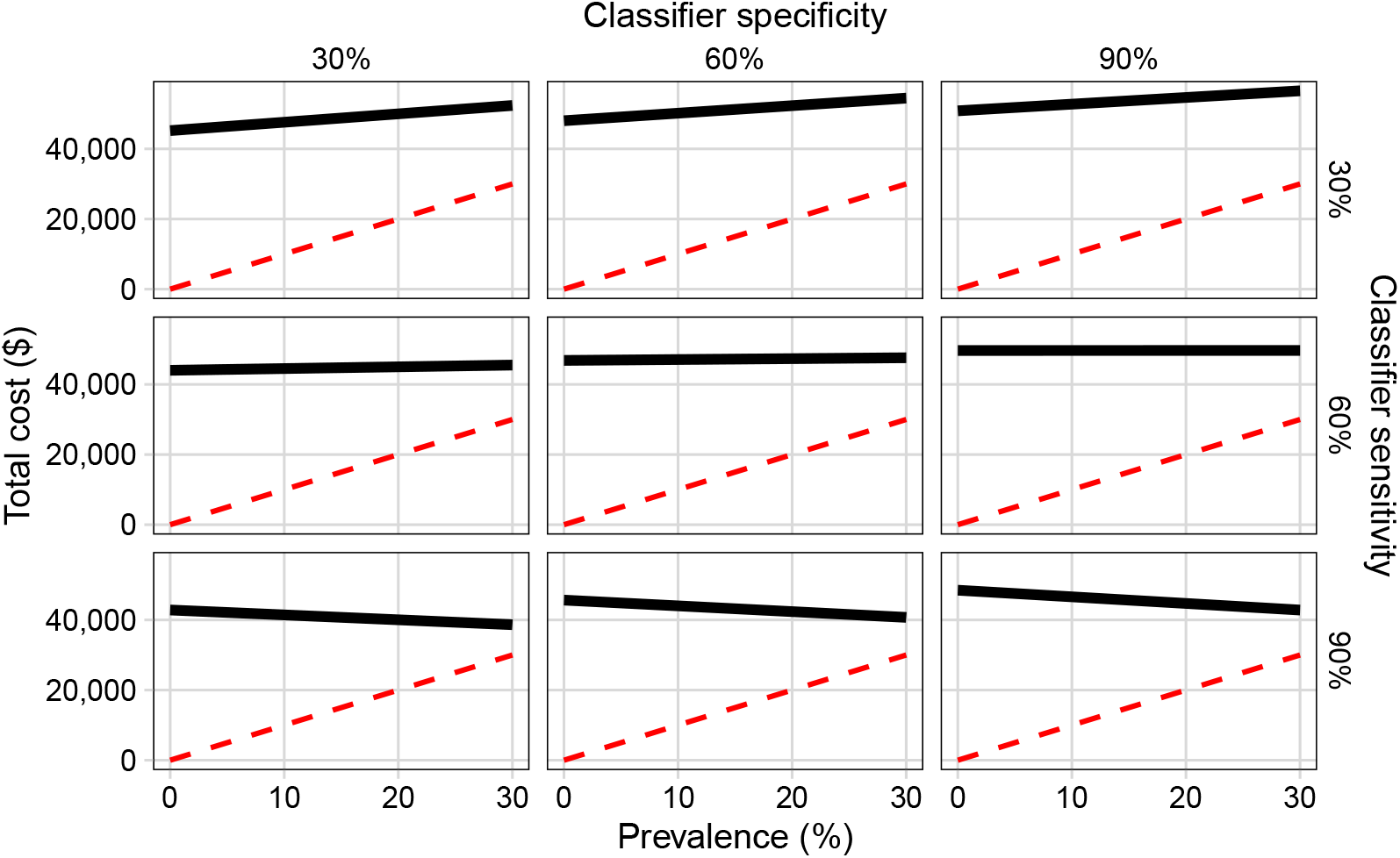
Total cost according to Strategy 4. Introducing RT-LAMP testing significantly decreases total costs compared with all other strategies.

### Positive Cases Reported

In the case of positive reported, Strategy 1 performs well in conjunction with a prior classification. Even when ignoring low-risk individuals, we capture almost all true positives when the sensitivity and specificity of the model is 90%. Sensitivity helps to discard potential true negatives, because it has determined correctly the majority of possible positive cases. When the sensitivity is low, the strategy misses those true positives who are thus left untested.

Strategy 2 includes the low-risk individuals (Figure 10), with an increase in positive reported from the start, decreasing the number of mismatches. Even when the classifier has low sensitivity and specificity, pooling captures the infected individuals identified as low-risk at the expense of higher costs than only using RT-qPCR.

**Fig 9.**
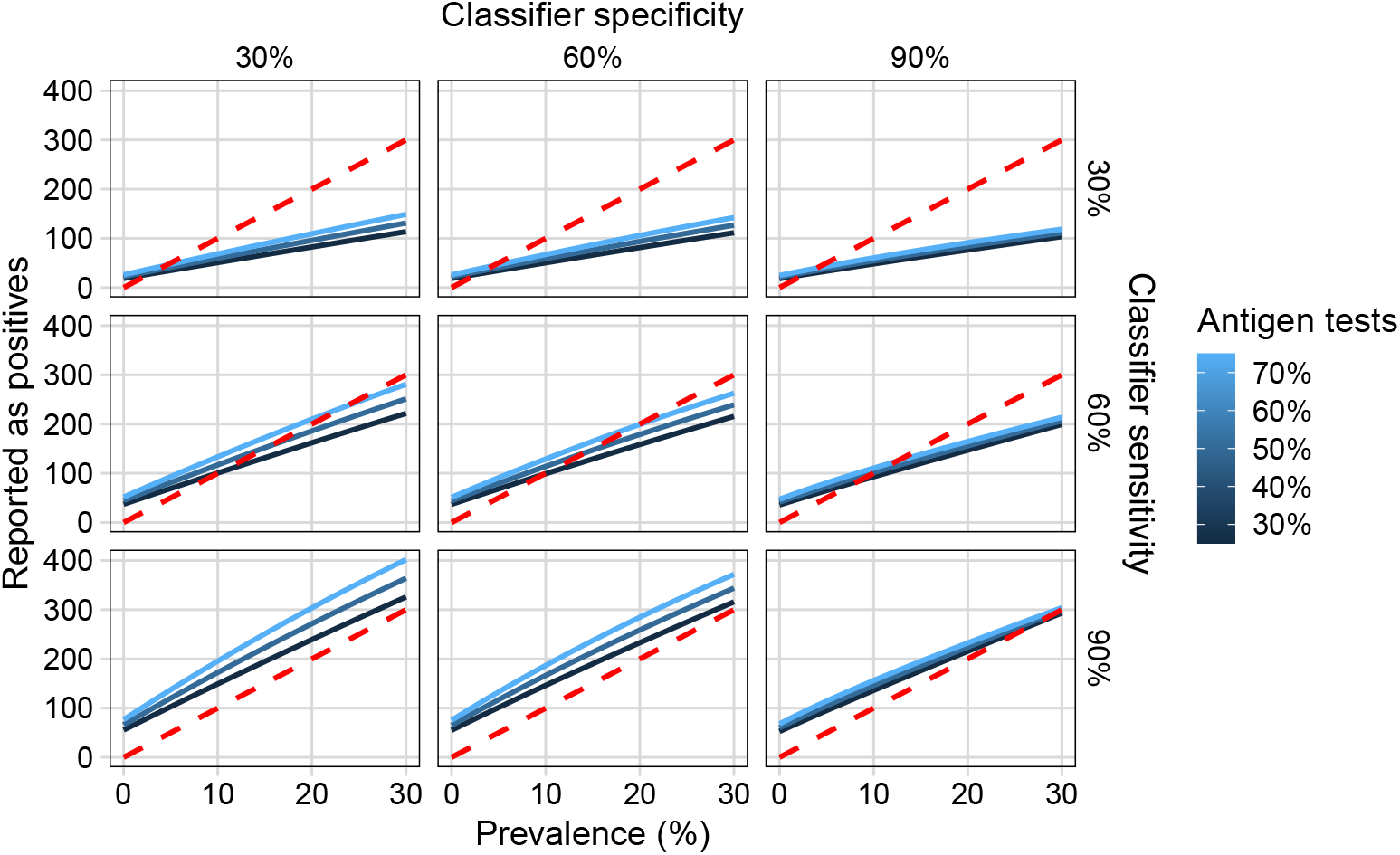
Number of individuals reported as positive according Strategy 1. Model sensitivity critically modulates detection of true positive cases.

**Fig 10.**
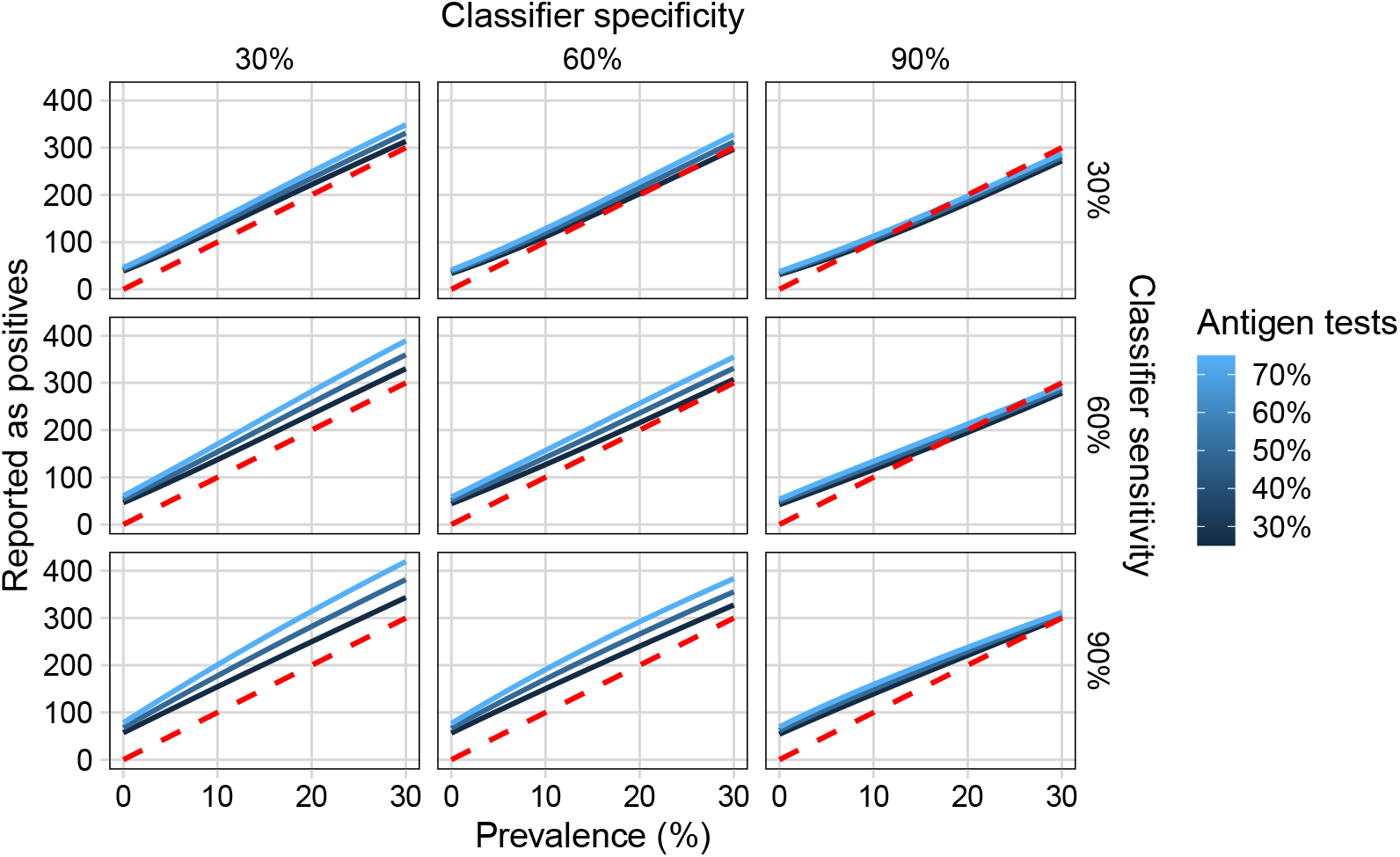
Number of individuals reported as positive according Strategy 2. Pooling improves detection at low prevalence compared to Strategy 1, observed in the smaller number of crossings between true cases and reported cases.

Strategy 3 (Figure 11) increase detection of true positives even more with respect to Strategy 1, specially at low prevalence contingent on reaching high sensitivity (90%); the number of false negatives increases at high prevalence below this sensitivity value. A large group of infected individuals are declared as low-risk. Combined with the application of antigen-based tests which have lower sensitivity than RT-qPCR ones, the probability of capturing true positives is reduced.

**Fig 11.**
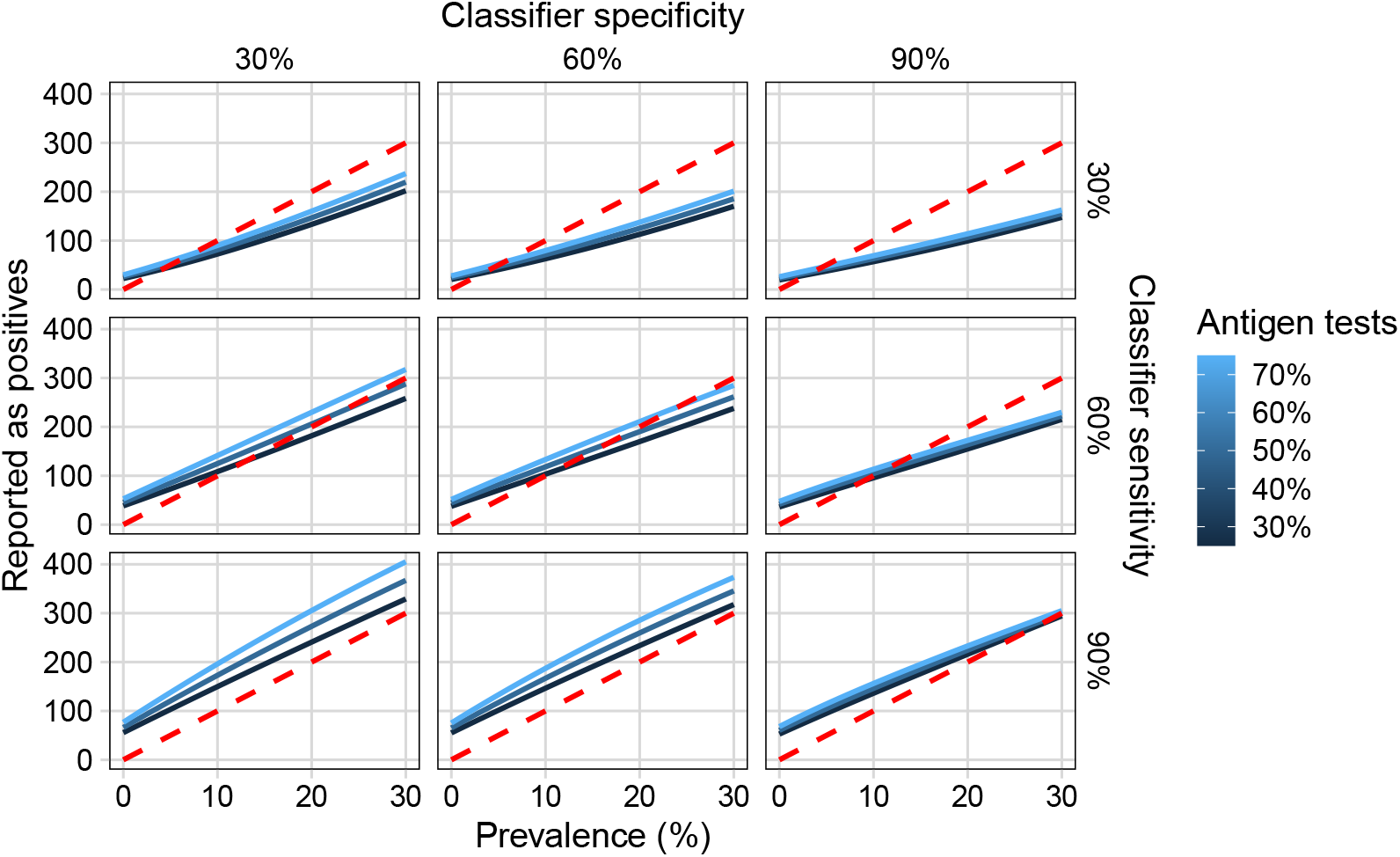
Number of reported as positive according Strategy 3. Sensitivity is inversely correlated with the number of false negatives as prevalence increases, explained in part by the lower sensitivity of antigen-based tests and the increasing reliance on them.

Strategy 4 (Figure 12) shows a similar pattern as Strategies 1 and 3. We observe that all functions are concave, implying improvements in detection as prevalence increases for sensitivity beyond 60%. Even when the outcome of the classifier resembles that of Strategies 1 and 3, the robustness of the curves indicates that RT-LAMP reduces the variability introduced by antigen-based testing.

**Fig 12.**
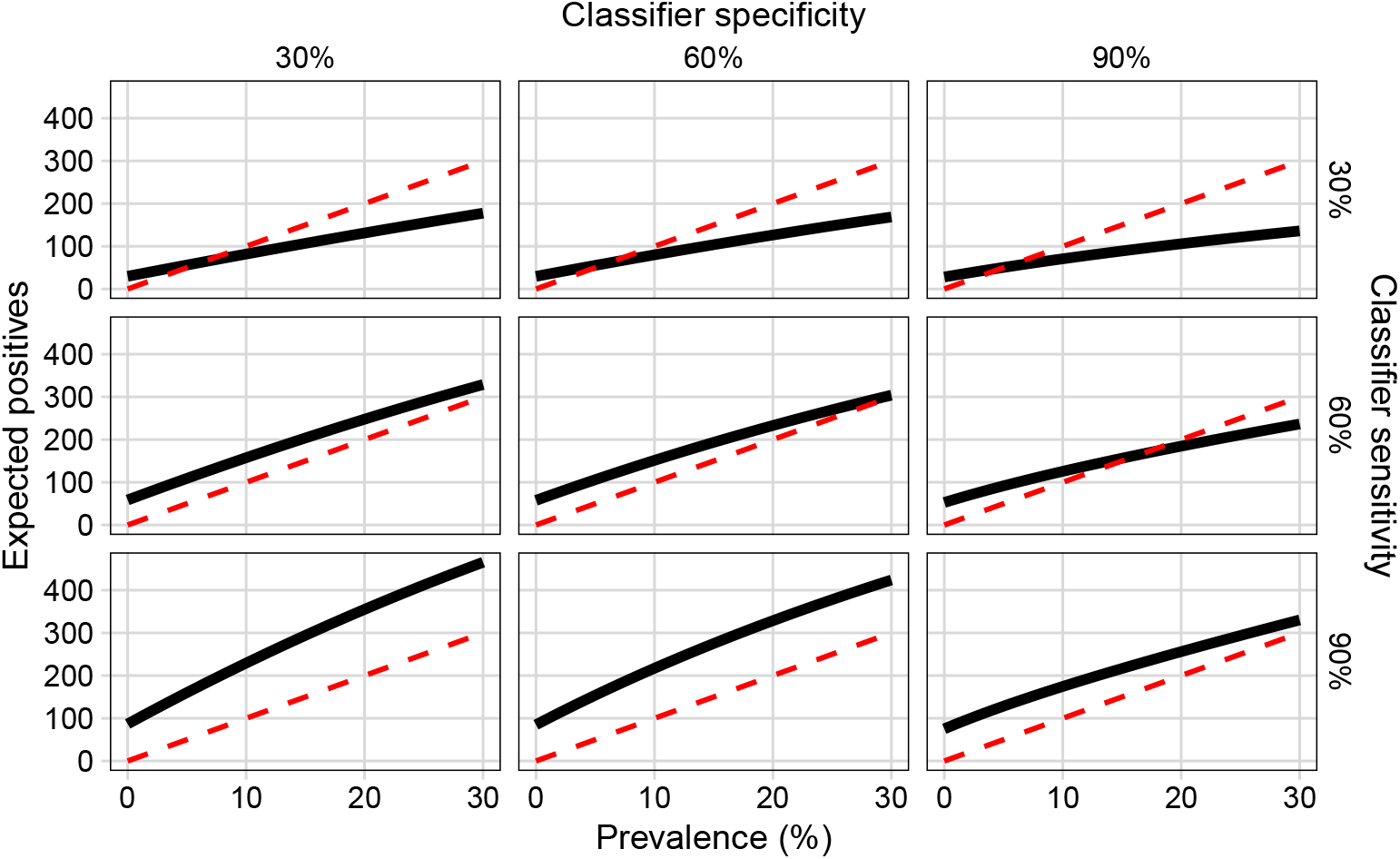
Number of reported as positive according Strategy 4. Introducing RT-LAMP testing yields concave curves at all sensitivity and specificity values. Outcomes are qualitatively similar to Strategies 1 and 3.

Finally, we observe that sensitivity below 50% appears to yield convex curves for number of positives reported, while curves corresponding to values above 50% seem to be all concave for strategies 1-3; this is modulated by the number of antigen-based tests when applicable. This is significant, since it delineates a response function in terms of testing efforts needed at a certain value of prevalence given a current combination of resources. The higher the prevalence, the more likely it is to increase detection of true positives. Similarly, the more antigen-based tests are used, the more likely false negatives will appear. However, it also implies that the impact of RT-LAMP and similar technologies is significant, since even at low sensitivity of the classifier the effort function is concave.

### Number of Tests per Person

For Strategy 1 (Figure 13), the number of tests per person obtained with computational experiments is as expected. The less accurate the model in identifying high-risk individuals, the larger the number of test needs to be be spent given the confirmatory mechanism of antigen-based testing against RT-qPCR. When the model is poorly fitted, the strategy spends around 1.2–1.7 tests per person. As the model sensitivity and specificity increases, the curves approach 1 at high prevalence. In all scenarios, the number of test per person is high (1.2–1.7) at low prevalence, since negatives are majority and the strategy must spend two tests to confirm true positives.

**Fig 13.**
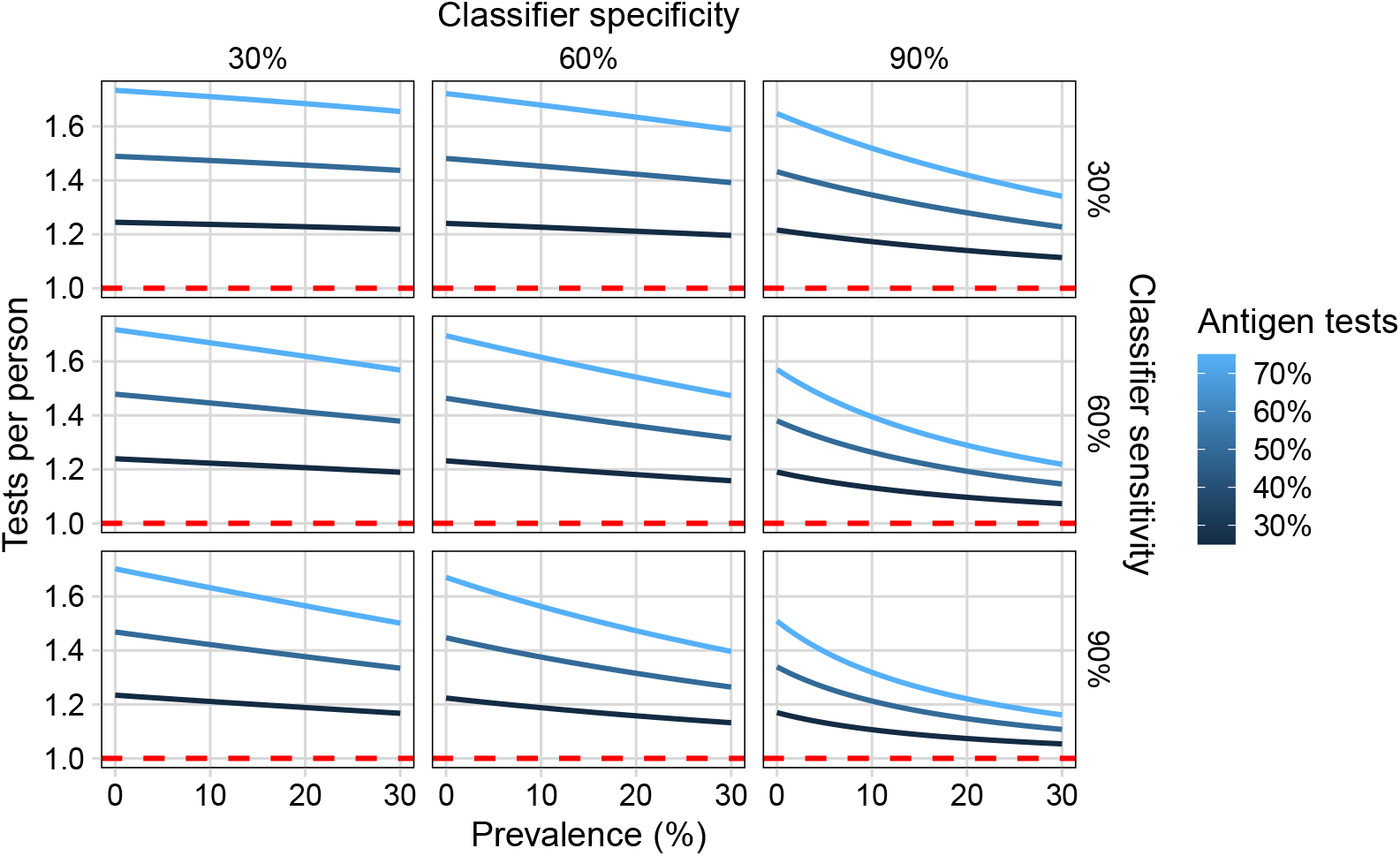
Number of tests per person in Strategy 1. Low prevalence forces more frequent retesting with antigen-based technologies, while high prevalence approximates one test per person at high sensitivity and specificity.

When pooling is introduced (Figure 14),a 0.5 reduction in average occurs when the model is correctly fitted with respect to Strategy 1. Specificity controls the behavior of the curve in terms of convexity and slope. Low specificity increases mis-classification of low-risk individuals, increasing the detection of true positives in the pooling technique.

**Fig 14.**
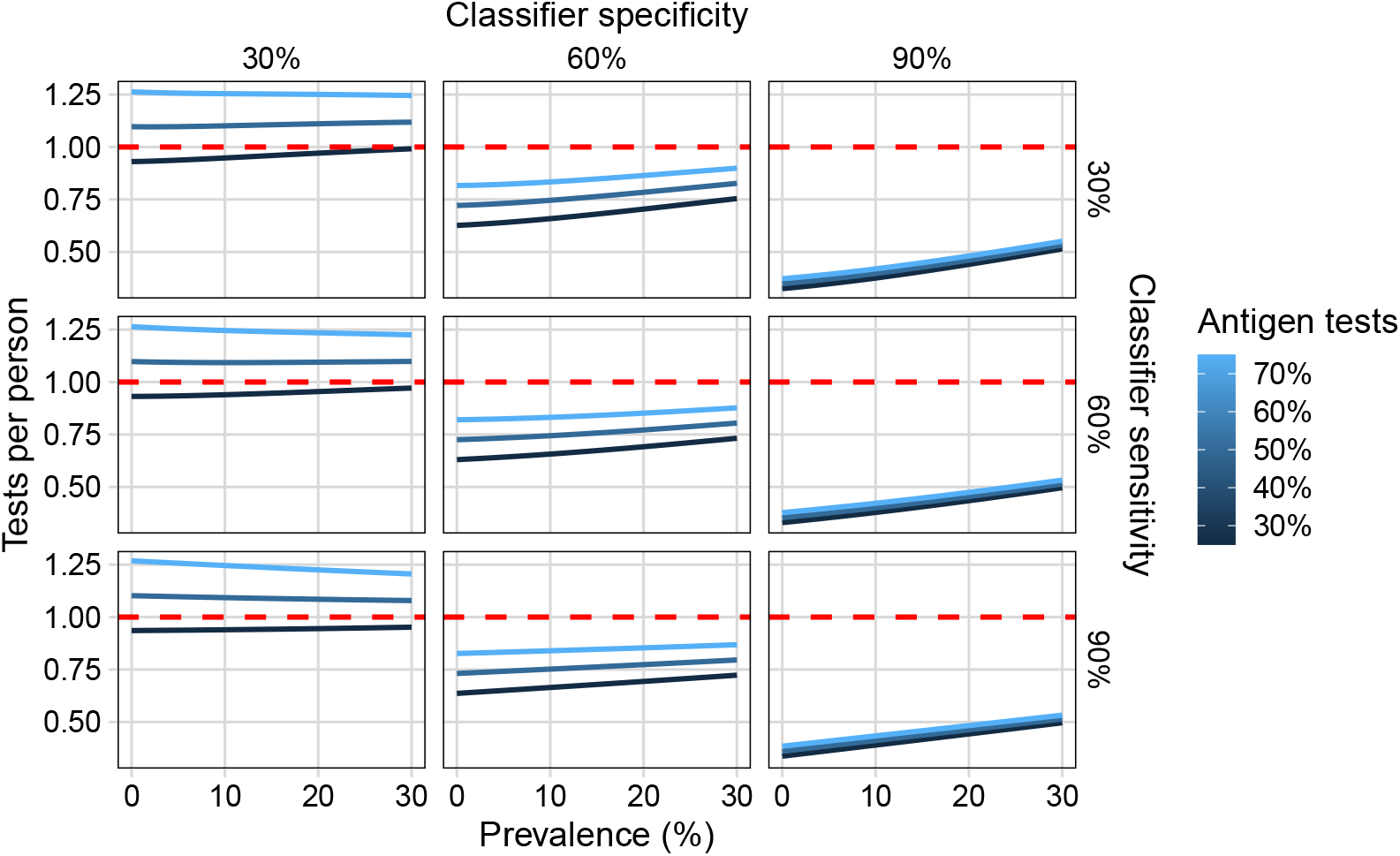
Number of tests per person in Strategy 2. An average of 0.5 fewer tests are needed, with a sharp decrease at high specificity, which controls the response of the system and erases differences introduced by sensitivity and the percentage of antigen-based tests applied.

The number of test per person in Strategy 3 descends linearly as specificity increases (Figure 15). Compared against Strategy 1, multiple testing can be reduced if the model is well-fitted. Strategy 2, in contrast, maintains better performance in this aspect. A similar pattern occurs in Strategy 4 (Figure 16). However, it is worth noting that the number of tests per person remains relatively constant –and close to 1-when the classifier shows high sensitivity and specificity in both Strategies. This is significant, since the resulting curve indicates scalability.

**Fig 15.**
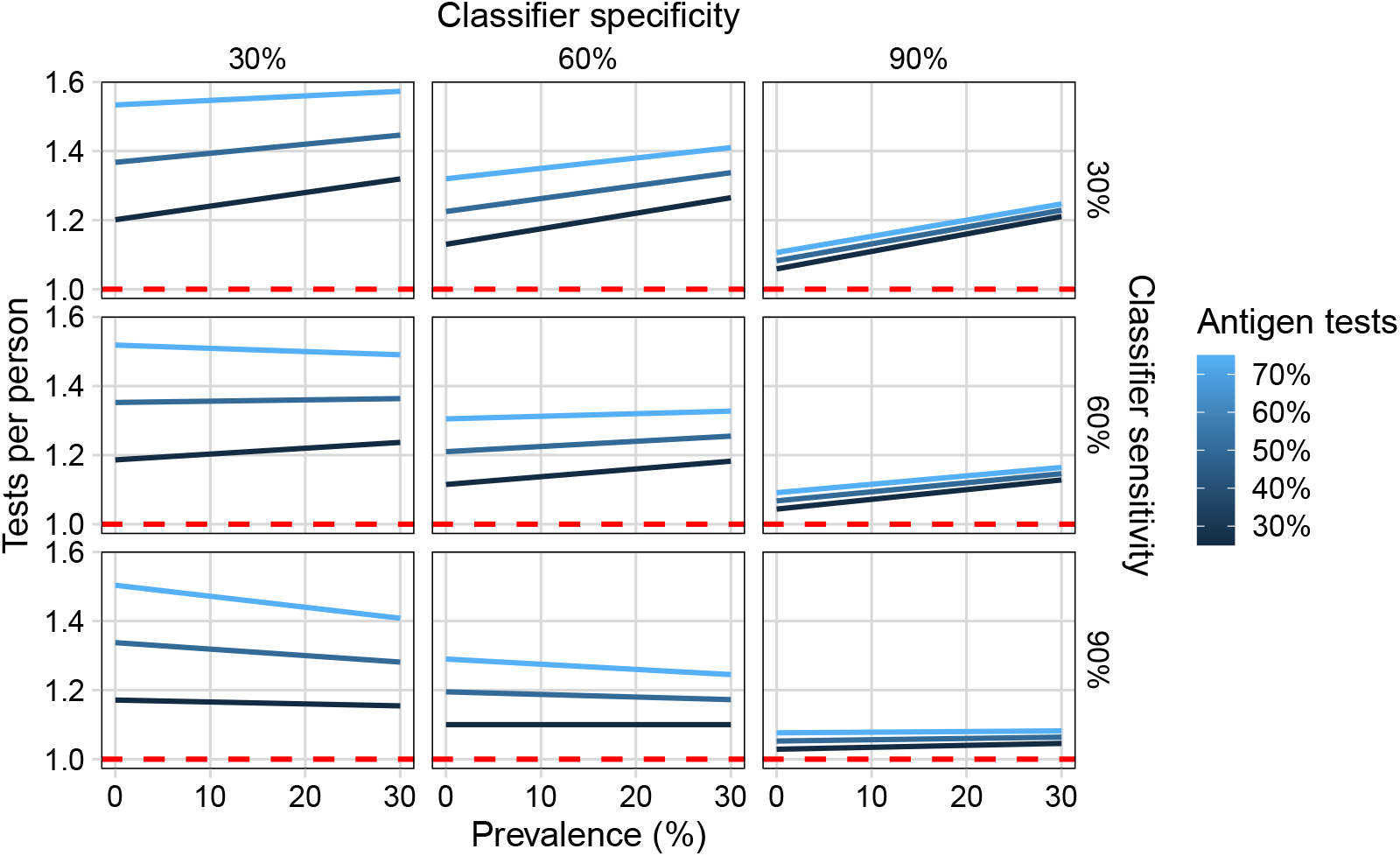
Test per person in Strategy 3. Specificity strongly determines proximity to one test per person, but cannot reach a a few tests as Strategy 2.

**Fig 16.**
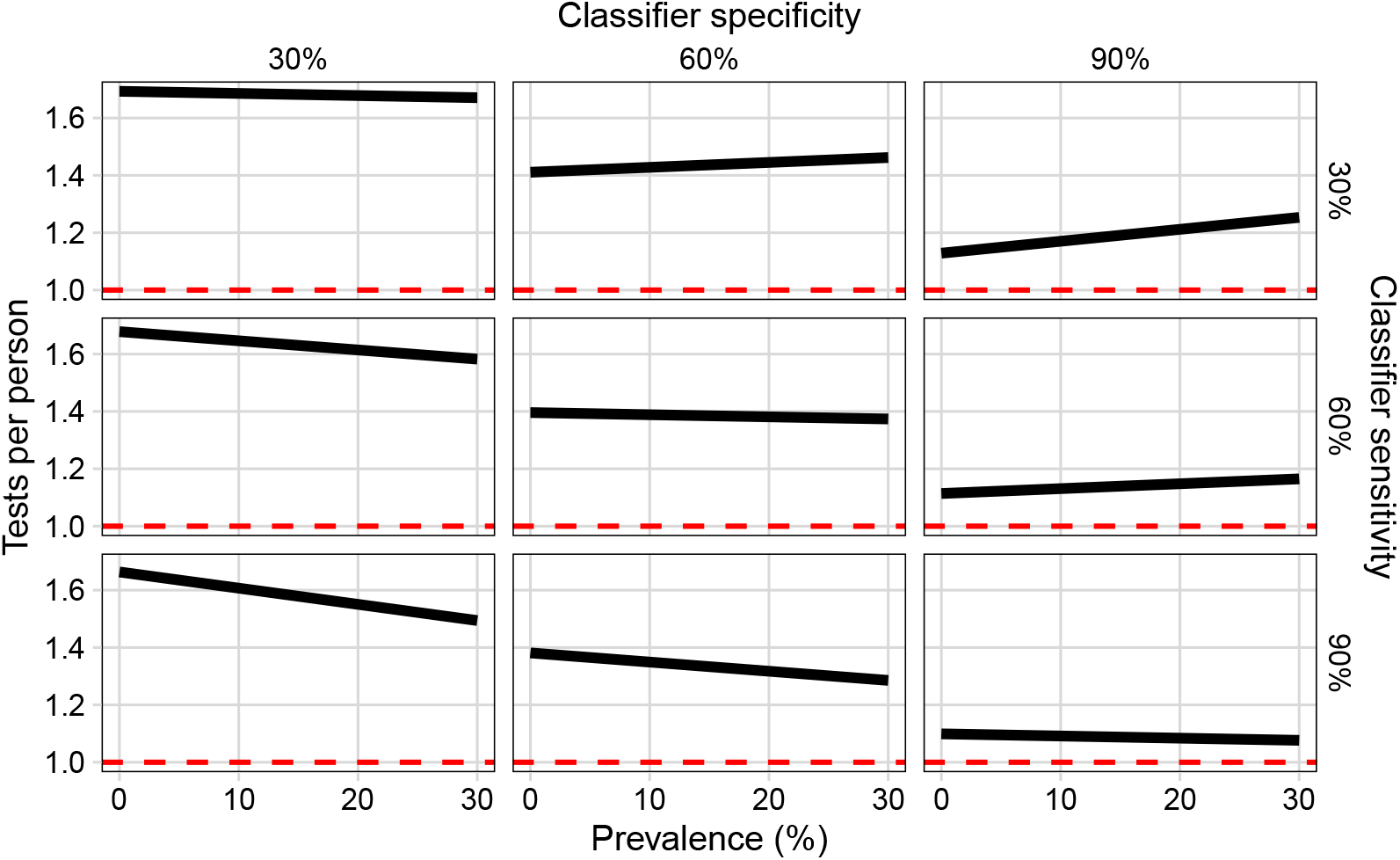
Test per person in Strategy 4. Similar to Strategy 3, specificity determines proximity to one test per person. The constant number of tests at high specificity and sensitivity, despite changes in prevalence, points to their scalability.

### Performance across strategies

To compare the relative performance across different strategies, we establish two new quantities, which we call *stock capacity* (*S*) and *detection efficiency* (*E*). To do so, we define an amortization index per Strategy *i*

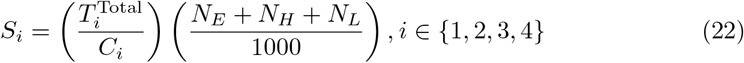

where 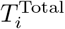 is the total number of test performed by that Strategy. The left-most factor in *S*_*i*_ represents the buying power of testing per each dollar spend. The right-most factor scales the number to the effectively covered population. This is the case of Strategy 1 where it only considers the high-risk population. For instance, if *S*_*i*_ = 0.01 and the budget is $100,000, then healthcare system can only afford *S*_*i*_ × 100,000 = 100 tests in total according to each strategy (a mix between RT-qPCR, Antigen and RT-LAMP).

Meanwhile, the detection efficiency is

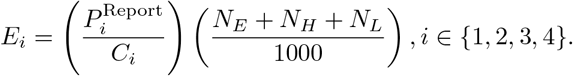

We interpret the index as the capacity of each strategy to detect a positive case per each dollar spend. Similar to *S*_*i*_ the number is scaled to the effective population covered. In the case of a value *E*_*i*_ = 0.001, and plans to spend $100,000 in the strategy, we can expect to detect *E*_*i*_ × $100,000 = 10 positive cases.

Figure 17 shows the values of *S*_*i*_ and *E*_*i*_ across all the strategies. We set here a fixed budget of $100,000. The red arrow (or point) represents a base case with detection of 1000 × ℙ(*D*_*P*_) positive cases spending $(100 × 1000 × ℙ(*D*_*P*_)) using only RT-qPCR tests. Arrows per strategy (i.e., hues of blue) indicate prevalence increase.

**Fig 17.**
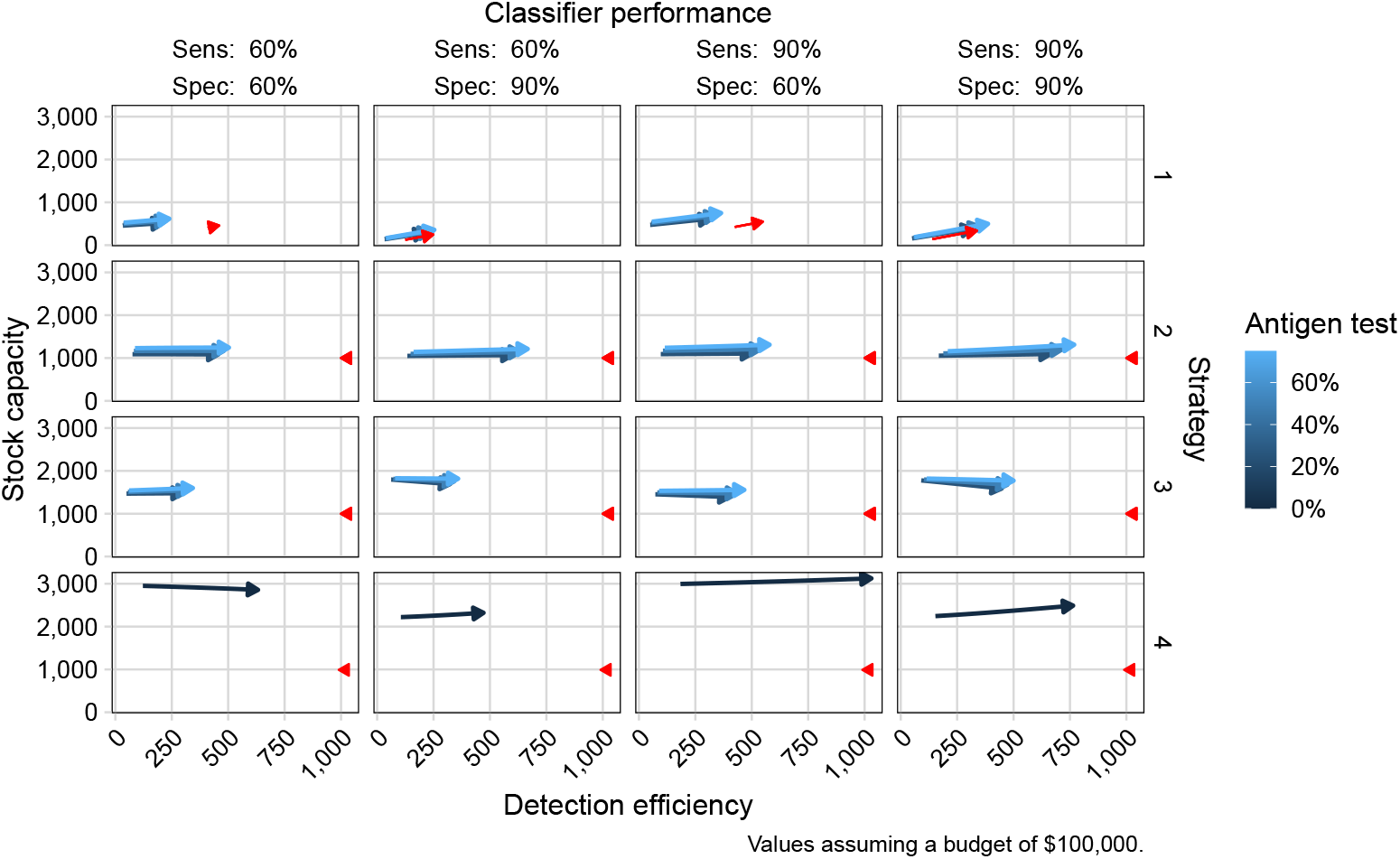
Comparison of the four strategies with respect to the stock capacity versus their detection efficiency. Arrows go from 0% (start) to 30% (point). Red arrows (or single triangle) represent the perfect case when there is 1000 people × ℙ(*D*_*P*_) positives, with a cost of 1000 people × ℙ(*D*_*P*_) × $100 using only RT-qPCR.

Strategy 1 shows its weakness due to the small capacity to buy tests and overall effectiveness. In other words, there is no difference between using antigen-based testing or one using only RT-qPCR if only patients classified as high-risk are tested in the best-case scenario, and significantly deteriorates when the classifier performs poorly. Meanwhile, Strategies 2 and 3 increase their capacities by covering the weak points from Strategy 1. Pooling (Strategy 2) increases the capacity of detection by maintaining the number of tests stable. Retesting (Strategy 3) is inefficient to capture positive cases even when the number of test is still small. This is explained due to the low sensitivity of antigen-based tests, around 80%.

Finally, Strategy 4 present an augmented buying power of tests and detection efficiency. Targeting technologies to specific pre-selected groups appears to be the best strategy to maximize budget impacts across healthcare systems.

## Discussion and conclusions

We studied in this article the theoretical impact of four different strategies for massive testing in Costa Rica in the case of COVID-19. We measured the overall cost of each strategy, the number of positive reported individuals and the number of test per person. To improve the performance of each strategy, we introduce a pre-classifier applied to the population before executing any possible testing campaigns. The putative classifier groups people into high and low risk, according to other variables like the social determinants of the population while preserving patient privacy and information security. Also, we presented a reformulation of the outcomes of each strategy in terms of purchasing power (i.e., stock capacity) detection effectiveness per dollar spend. Our theoretical analysis provides a better picture of the impact of different strategies for massive testing on performance and resource allocation during a pandemic. While vaccination has become widely available, remaining vigilant of the phenomenon and preparing for massive testing is crucial for containing any new variants or other infectious diseases.

Our results show that Strategy 1 can be improved only by adequately identifying true positive individuals, while ignoring altogether the low-risk group; this makes it indistinguishable to not just applying routine RT-qPCR tests as usual. Strategy 2 exhibits better testing coverage by including the pooling technique for low-risk individuals. The technique however, is rendered ineffective when a large group of potential positives are tested; the technique requires more testing than just applying one test per person. Strategy 3 is more straightforward to implement. We discovered, however, that it is more costly due to multiple testing in the low-risk group. Finally, Strategy 4 is the most cost-effective strategy due to both the properties of the testing technology (RT-LAMP) and the more refined targeting of the testing protocol per risk group.

The introduction of a predictive model or classifier brings two strategic advantages. First, it can reduce overall costs, time and human efforts. Second, it increases information richness across the testing process. The first advantage relates to the system capacity to choose the best and cheaper technology according to each patient. If the model classifies individuals correctly, testing efforts can be optimized. Furthermore, healthcare systems can cover deficiencies present in one technology with the advantages of another (i.e., scalability), using the probabilistic prediction of the classifier as a triaging device while waiting for laboratory tests to finish and confirm or reject the result. Having more data, and consequently better prediction capabilities, allows clustering individuals into subgroups according to particular features such as their social, demographic or economic indicators and mobility patterns, among others. This information could lead healthcare authorities to adopt more personalized measures to cover certain vulnerable groups.

Our results show that all the strategies become more effective when the classifier –arguably a sophisticated machine learning method-is well-fitted, reaching sensitivity and specificity levels of 60% or higher. In particular, we showed that sensitivity (identification of potential positives) plays a crucial role in reducing costs and increasing confirmation of positives. For the pooling scenario, specificity controls the number of tests per person.

One of the fundamental limitations of achieving a good fit for such models is access to high-quality individual data. The quality of data remains a challenge since the beginning of the pandemic, particularly in developing nations and emerging economies. Available data tend to only reflect the reality of people who have undergone testing, and even when that is the case, datasets are biased by the administrative reality –and shortcomings-of the specific healthcare system. Therefore, we can expect a similar systematic bias in the classification process due to the different epidemiological moments across the pandemic. Testing increases during high-peak waves, confirming symptomatic patients and capturing asymptomatic nexus of them. When the pandemic wave passes and minimum cases are reached, the testing strategy tends to focus on confirming symptomatic cases arriving at clinical centers. During these periods, the real number of infected asymptomatic people remains unclear. In addition, overloading of the healthcare services impacts data production, which may be ready for consumption days or weeks later. This requires, as proposed, adjusting the model to correct for administrative and systematic lags.

Another set of limitations corresponds to the choice of potential classification models as well. We mention a non-exhaustive list of classification methods with their respective advantages and disadvantages. The classic logistic regression model is easy to implement, but the implicit assumptions and the inclusion of administrative lags in the data can negatively impact the interpretability of results due to an artificial increase in the number of coefficients; in this situation, a Ridge or Lasso regularization could reduce their number. Another option, if the data exhibits non-linearity, is to use a support vector machine (SVM), which can handle situations in which classes are not-linearly separable. The downside here is the computational cost during the training stage, which has to be performed a limited number of times as the pandemic evolves. Tree ensemble approaches are popular, including Random Forest, XGboost, and Gradient Boosting. In practice, these methods perform better than the mentioned classifiers, but require fine-tuning of hyperparameters whose interpretation may not be direct. Finally, deep-learning algorithms can be used to fit the classifier at the expense of complexity and interpretability.

We envision a series of challenges in the implementation of a classification system such as that described here. The main one is the adoption of machine learning assisted system by clinical and health policy authorities to triage the population before performing laboratory tests. While unforeseen clinical or ethical reasons may hamper the implementation of the model, the aim of this statistical approach is to become a companion instead of competitor for healthcare providers. The advantage of classification-assisted triaging of patients in clinical contexts has been discussed and demonstrated in literature in general [74, 75], and more recently in the context of the COVID-19 pandemic [76, 77]. Having some prior information about the possible test result can better prepare clinicians and staff to handling wave peaks efficiently, allocate resources more appropriately and anticipate critical resource usage and patient mortality counterfactuals. Another challenge is the actual capacity of systems triage patients. Even with an algorithm ready, further studies are needed about how to integrate it into workflows across medical centers and public health authorities. In the particular case of Costa Rica, the EDUS (*Expediente Digital Único en Salud*) system can serve as the the channel to deliver results from the algorithm to laboratory technicians and physicians. However, creation of a new submodule will require testing, validation and data assurance in compliance with information security standards in the public health service (CCSS). Even if the EDUS system already collects already most of the information about patients, the process of anonymizing, handling, securing, and ensuring responsible use of personal information must remain as a top priority. Finally, the attitude of the public around collection of information and its handling constitutes a challenge of uncertain proportions.

Our next step is to fit a classifier using both real and synthetic datasets. The EDUS is the main source of individual data of the Costa Rican public health. When a patient arrives at a medical appointment, physicians register the health status, diagnosis, demographic and related factors of each patient. During the COVID-19 pandemic, the tool was used to track down the symptoms across the population, to provide hot-lines for medical support and to validate the number of vaccines already applied. We believe this information source can be responsibly used further in benefit of all users. Its main advantage is the massive information density and patient coverage. Given the universal healthcare system in Costa Rica, information about a wide range of groups exists regardless of economic status. Another secondary corresponds to the *Instituto Costarricense de Investigación y Enseñanza en Nutrición y Salud* (INCIENSA: National Institute of Research and Education on Nutrition and Health). At the beginning of the pandemic, INCIENSA collected numerous COVID-19samples alongside epidemiological and sociodemographic data of infected patients. Even if the diversity in this source is less than that of EDUS, it could be an important source to adjust the model.

Finally, we expect to develop synthetic datasets through simulation. Prior experience with agent-based modeling [17] indicate that it is possible to replicate features of epidemic waves and the effect of public policy measures *in silico*, to then overlay our strategies and determine performance under various scenarios and constraints; other methods exist, and will be explored. These datasets can be openly shared across all relevant stakeholders without risking healthcare data leaks while still being representative of aggregate statistics of the underlying population.

## Data Availability

All data produced are available online at https://github.com/maikol-solis/code_paper_massive_testing_strategies

## Acknowledgments

This project was supported by the TICOVID project at the University of Costa Rica (internal grant number 817-C0530-22), the Embassy of France in Costa Rica, and the National Center for Supercomputing Applications at the University of Illinois Urbana-Champaign. Special thanks to Dr. Andrés Gatica-Arias and Dr. Germán Madrigal-Redondo for creating the TICOVID project and promoting the transdisciplinary research from the beginning. We thank to them and other TICOVID project for their encouragement and input on COVID-19 specific questions on testing technologies.

## Footnotes

1 See: https://github.com/maikol-solis/code_paper_massive_testing_strategies

